# Germ cell-specific proteins ACRV1 and AKAP4 facilitate identification of rare spermatozoa in semen of non-obstructive azoospermia patients

**DOI:** 10.1101/2022.09.15.22280011

**Authors:** Junyan Zhang, Mirzo Kanoatov, Keith Jarvi, Andrée Gauthier-Fisher, Sergey I. Moskovtsev, Clifford Librach, Andrei P. Drabovich

## Abstract

Non-obstructive azoospermia (NOA), the most severe form of male infertility due to testicular failure, could be treated with intra-cytoplasmic sperm injection (ICSI), providing spermatozoa were retrieved with the microdissection testicular sperm extraction (mTESE). Here, we hypothesized that some testis- and germ cell-specific proteins would facilitate flow cytometry-assisted identification of rare spermatozoa in semen cell pellets of NOA patients, thus enabling non-invasive diagnostics prior to mTESE. Data mining and extensive verification by targeted proteomic assays and immunofluorescent microscopy revealed a panel of testis-specific proteins expressed at the continuum of germ cell differentiation, including the late germ cell-specific proteins AKAP4_HUMAN and ASPX_HUMAN (ACRV1 gene) with the exclusive expression in spermatozoa tails and acrosomes, respectively. A multiplex imaging flow cytometry assay revealed low numbers of the morphologically intact AKAP4^+^/ASPX^+^/Hoechst^+^ spermatozoa in semen pellet of NOA patients. While the previously suggested soluble markers for spermatozoa retrieval suffered from low diagnostic specificity, our multi-step gating strategy and visualization of AKAP4^+^/ASPX^+^/Hoechst^+^ cells bearing elongated tails and acrosome-capped nuclei facilitated fast and unambiguous identification of the mature intact spermatozoa. Pending further validation, our assay may emerge as a non-invasive test to predict the retrieval of morphologically intact spermatozoa by mTESE, thus improving diagnostics and treatment of the severe forms of male infertility.

## INTRODUCTION

Infertility affects 15% of couples globally, with a male factor contributing to the infertility in approximately 50% of the cases *(1)*. The routine diagnostics of male factor infertility relies on semen analysis for the spermatozoa count, motility and morphology, and on infertility in the couple *(2)*. Common categories of male infertility based on abnormal sperm parameters include oligozoospermia, asthenozoospermia, teratozoospermia, azoospermia, and a combination of morphological abnormalities with low sperm count and diminished motility diagnosed as oligoasthenoteratozoospermia. Azoospermia is further classified as obstructive azoospermia (OA) with no sperm in ejaculate despite normal spermatogenesis, and non-obstructive azoospermia (NOA) with no sperm in ejaculate due to testicular failure *(2)*. Men with elevated follicle-stimulating hormone (FSH) in blood serum and smaller testicles are more likely to have testicular failure resulting in NOA. However, men with normal FSH levels could have either NOA or OA *(3)*. A testicular biopsy to determine the presence and degree of spermatogenesis is typically required to distinguish OA and NOA.

Breakthroughs in assisted reproductive technologies allowed fertility clinics to offer successful treatments to couples with male factor infertility, including its severe forms *(4)*. Patients with mild oligozoospermia or idiopathic male infertility with normal semen parameters have sufficient numbers of spermatozoa in the semen to be offered either intra-uterine insemination (IUI), conventional *in-vitro* fertilization (IVF), or intra-cytoplasmic sperm injection (ICSI). Patients with moderate oligoasthenoteratozoospermia could be managed with IVF or ICSI depending on sperm count and spermatozoa quality. NOA, the most severe form of male infertility, could only be treated with ICSI, providing spermatozoa could be retrieved through microdissection testicular sperm extraction (mTESE). MTESE has ∼50% success rate and can identify spermatozoa even in those cases when initial testicular biopsy revealed no spermatozoa *(5, 6)*. It should be noted that diagnostic testicular needle biopsies, partially due to the random sampling, fail to identify spermatozoa in ∼30% NOA patients which had spermatozoa following mTESE *(7)*. Thus, extensive microsurgical dissection of seminiferous tubules and their examination with an operating microscope is the most widely used technique to identify pockets of spermatozoa. Seminiferous tubules with spermatogenesis are excised, and the mTESE spermatozoa are used for ICSI, with the reported pregnancy rates between 20–50% *(8, 9)*. The mTESE procedure is laborious, and patients often spend more than three hours under general anesthesia. The heterogeneous etiology of male infertility, low diagnostic sensitivity of FSH, inaccuracy of the initial testicular biopsy diagnostics, and laborious surgical retrieval of spermatozoa may result in different and unpredictable mTESE outcomes at different fertility centers. A non-invasive diagnostic test to predict mTESE outcomes with high specificity constitutes a recognized clinical need. Likewise, a viable approach for the rapid identification of rare mature spermatozoa within dissected seminiferous tubules of NOA patients, or within semen cell debris of patients with severe oligoasthenoteratozoospermia, would be highly beneficial to IVF programs.

There were numbers of studies on the identification of novel biomarkers of male infertility, including genomic *(10-12)* and epigenetic biomarkers *(13)*, RNA *(14)*, metabolites *(15)*, hormones *(16)*, and proteins *(17-22)*. Numerous parameters including testis size, genetic profiles, serum testosterone, FSH, inhibin B, and seminal proteins were evaluated to predict mTESE sperm retrieval outcome *(23, 24)*. Likewise, we recently developed a non-invasive test to differentiate between NOA and OA *(25, 26)*. While those approaches revealed diagnostic sensitivity of 60-80%, they suffered from low specificity *(27-29)*. Interestingly, low diagnostic specificity for mTESE retrieval of spermatozoa was found even for the highly promising testis-specific proteins, likely due to their expression at the continuum of germ cell differentiation and presence at the early and late stages of spermatogenesis *(28)*.

In this study, we hypothesized that testis-specific proteins further categorized as germ cell-specific could facilitate more specific and non-invasive identification of rare spermatozoa in the NOA semen cell pellets by flow cytometry. In addition, we suggested that imaging flow cytometry with multiplex visualization of the organelle-specific proteins could also reveal spermatozoa integrity. The unique morphology of spermatozoa including acrosome-capped nuclei and elongated tails could be visualized with the aid of several testis-, germ cell- and organelle-specific markers and could provide unambiguous identification of spermatozoa with nearly no false-positives and high diagnostic specificity unachievable with the previously reported soluble biomarkers in semen.

Our pipeline to identify the most promising candidate markers relied on data mining for the testis and germ cell-specific proteins, in-depth identification and quantification of the spermatozoa proteome, extensive validation by the targeted proteomic assays, and selection of protein markers which could be visualized with the multiplex immunofluorescent microscopy and imaging flow cytometry assays.

## RESULTS

### Data mining and identification of testis- and germ cell-specific proteins

We previously identified one of the largest proteomes of human spermatozoa (8,046 proteins) *(30)* and seminal plasma (SP; 3,141 proteins) *(31-33)*. A combined proteome was searched across the Human Protein Atlas *(34)* and NextProt *(35)* data and revealed 578 testis-specific proteins expressed in spermatozoa (**Fig. 1A** and **supplemental table S1**). Proteins with at least one extracellular membrane-bound isoform (129 proteins) or a secreted isoform (55 proteins) were prioritized as the most promising markers. We also suggested that some proteins predicted as exclusively membrane-bound could have non-canonical secreted or membrane-shed isoforms, and vice versa. For example, secreted acrosomal protein ASPX_HUMAN, the product of ACRV1 gene with 11 different transcript isoforms, was previously found as a soluble protein in the acrosomal matrix and as a protein bound to the inner acrosomal membrane *(36)*. Human Protein Atlas immunohistochemistry data for the membrane-bound and secreted proteins were manually examined to select 45 proteins expressed at the different stages of spermatogenesis (**Fig. 1B** and **table S2**), excluding mature spermatozoa due to low 40× magnification. Finally, Antibodypedia portal *(37)* was searched for immunoassays and antibodies suitable to develop experimental validation assays, but revealed that the high-quality antibodies were not available for the majority of selected testis-specific proteins. Thus, we proposed targeted proteomic assays as tools to experimentally evaluate the abundance of candidate proteins in spermatozoa, SP and vas deferens fluid samples.

**Fig. 1.**
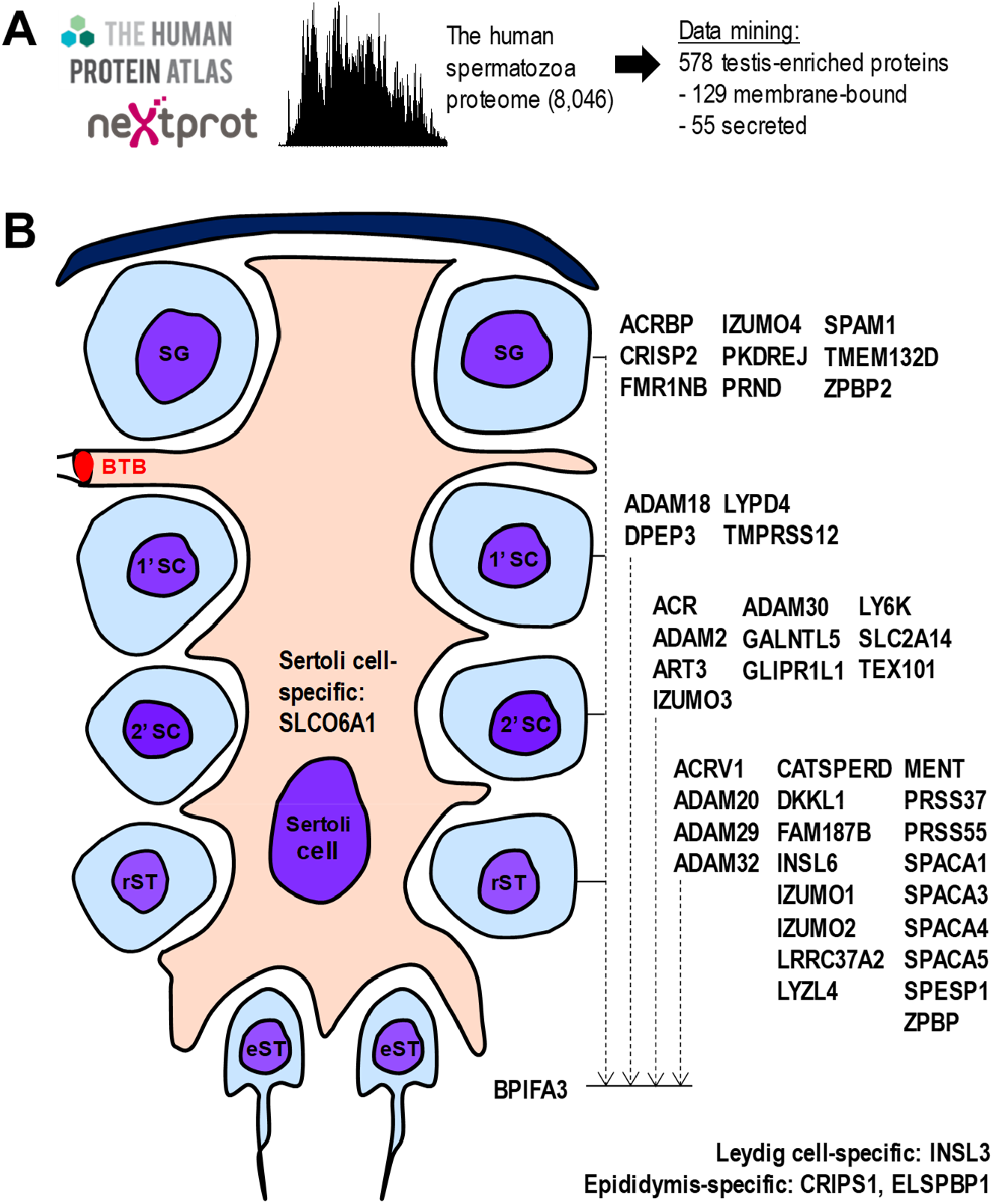
Data mining to select testis germ cell-specific proteins. (**A**) Deep proteome of the human spermatozoa *(30)*, Human Protein Atlas and NextProt data were used to identify and classify testis-enriched, membrane-bound and secreted genes and proteins. (**B**) Human Protein Atlas immunohistochemistry data was manually examined to select genes and proteins expressed at the continuum of germ cell differentiation, excluding spermatozoa. Human gene names are presented. BTB, blood-testis barrier; SG, spermatogonia; 1’SC, preleptotene spermatocytes; 2’SC, pachytene spermatocytes; rST, round spermatids; eST, elongated spermatids.

### Development of multiplex quantitative targeted proteomic assays

We previously demonstrated the application of targeted proteomic assays for protein quantification in various biological fluids and clinical samples *(38-41)*. To develop SRM assays, we re-searched our previous shotgun mass spectrometry and label-free quantification data on protein expression in human spermatozoa and SP *(30, 33)*, and selected the unique and high-intensity proteotypic peptides and their most intense y- and b-ion fragments. Following several rounds of assay development and peptide prioritization, the most promising proteotypic peptides were synthesized as heavy isotope-labeled peptides and used as internal standards for development of quantitative targeted proteomics assays (**tables S3-4**).

### Quantification of testis-specific proteins in spermatozoa, vas deferens fluid and SP

We hypothesized that the most promising protein markers to detect mature spermatozoa would be (i) abundant in spermatozoa lysate; (ii) reproducibly measured in vas deferens fluid and SP suggesting their endogenous shedding from the cell surface or secretion, and potentially excluding intracellular proteins leaking out of the damaged spermatozoa. We quantified by SRM the abundance of 45 germ cell-specific proteins in normozoospermic spermatozoa, vas deferens fluid and SP samples (**Fig. 2A**) and selected 18 most promising proteins (**Table 1**). We further optimized our multiplex SRM assay (**table S5**) and measured the levels of 18 testis-specific proteins in matched pre- and post-vasectomy SP samples (**fig. S1**). As a result, levels of twelve proteins decreased below the SRM assay limits of detection, suggesting high testis tissue specificity. To provide the more detailed evaluation, 18 proteins were then measured in the larger independent sets of SP samples including unmatched pre- and post-vasectomy, OA and NOA with the known histological subtypes of hypospermatogenesis, maturation arrest and Sertoli cell-only (**fig. S2**). Interestingly, the results re-discovered TX101_HUMAN and DPEP3_HUMAN proteins reported in our previous studies *(25, 42)* and re-confirmed their diagnostic performance to non-invasively detect post-vasectomy and OA with AUC=1.0. In addition, we identified a superior protein ASPX_HUMAN (acrosomal protein SP-10; ACRV1 gene) which levels decreased 61-fold in post-vasectomy SP and provided AUC=1.0 at its limit of detection of 0.4 μg/mL. As a result, we focused on ASPX_HUMAN as the most promising soluble marker for the non-invasive diagnosis of post-vasectomy, OA and NOA (**Table 1** and **Fig. 2B-C**). It should also be noted that two epididymis-specific proteins ESPB1_HUMAN and CRIS1_HUMAN emerged as novel markers to differentiate between NOA and OA/PV. ESPB1_HUMAN (AUC=0.72, MWU *P*=8×10^−4^) and CRIS1_HUMAN (AUC=0.73, MWU *P*=6×10^−4^) could be useful to complement ECM1_HUMAN for the differential diagnosis of azoospermia, as we previously reported *(25, 28)*.

**Table 1.**
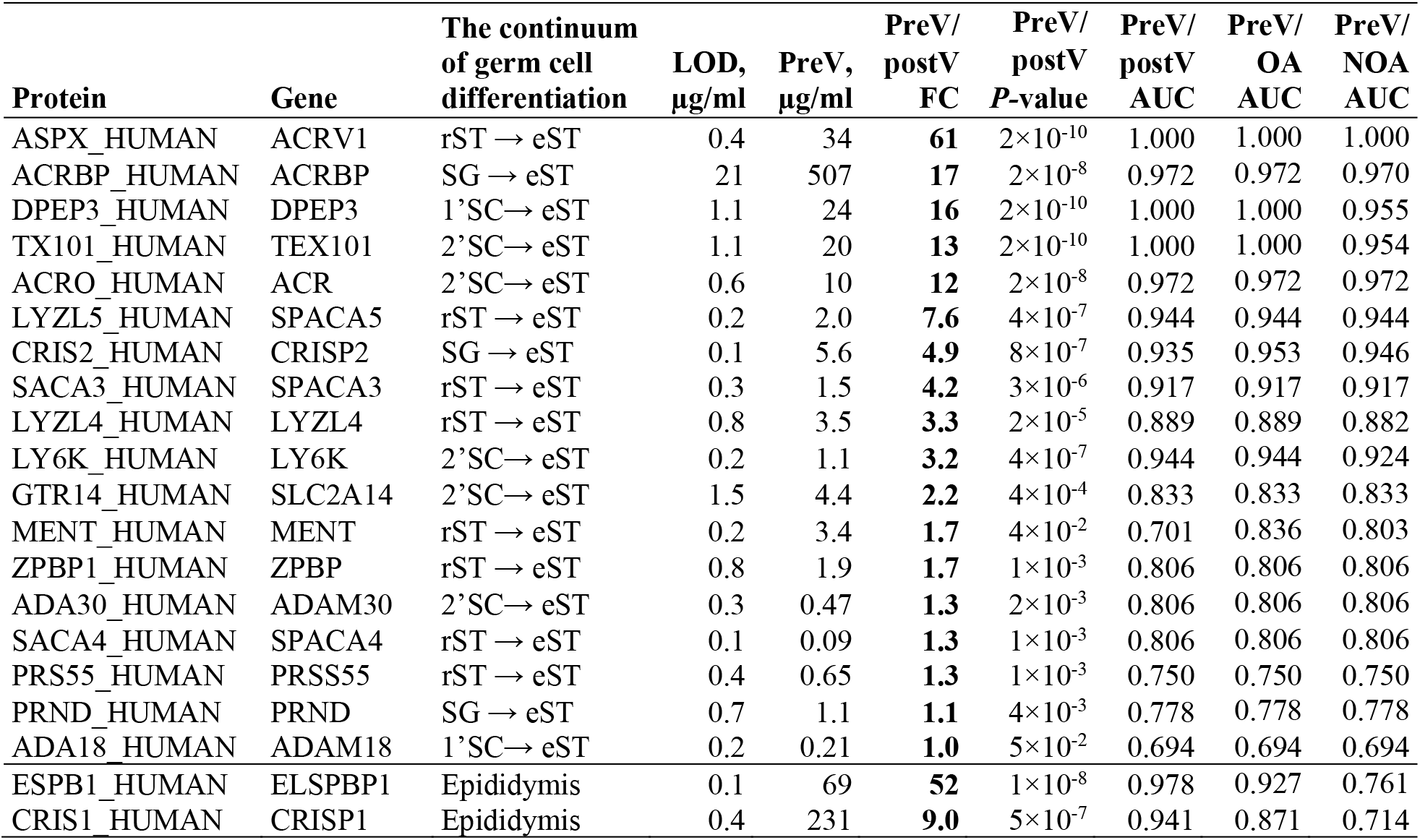
Diagnostic performance of 18 testis germ cell-specific proteins measured by SRM assays in SP. The continuum of germ cell specificity (spermatogonia to elongated spermatids, excluding mature spermatozoa) was provided by the Human Protein Atlas immunohistochemistry data. Epididymis-specific proteins were measured as controls. SP cohorts included normozoospermic pre-vasectomy (preV), unmatched post-vasectomy (postV), OA and NOA samples. SRM assay limits of detection (LOD) were calculated using serial dilutions of heavy peptides spiked into the digest of preV SP pool and measured within the linear response range of H/L ratio of calibration curves. To calculate fold changes of median concentrations (FC), two-tailed Mann–Whitney U test *P-*values and areas under the receiver operating characteristic curve (AUCs), the levels below LOD were adjusted to LOD. SG, spermatogonia; 1’SC, preleptotene spermatocytes; 2’SC, pachytene spermatocytes; rST, round spermatids; eST, elongated spermatids.

**Fig. 2.**
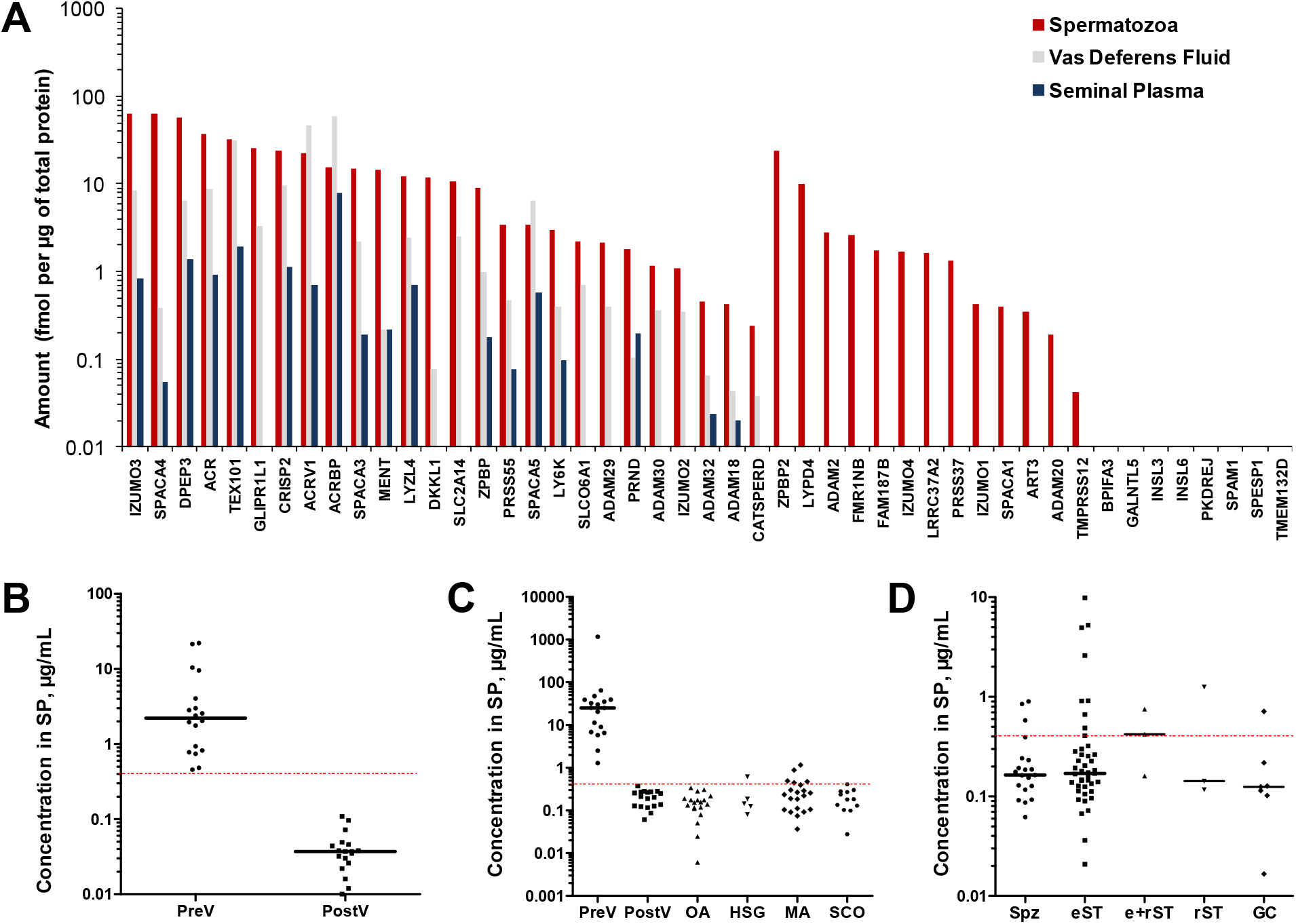
Levels of testis germ cell-specific proteins in human spermatozoa, vas deferens fluid and SP measured by SRM assays. **(A)** 45 germ cell-specific proteins were measured in a pool of spermatozoa lysates and matched SP from the fertile normozoospermic men, as well as unmatched vas deferens fluid samples collected during vasectomy. A Sertoli cell-specific SLCO6A1 and Leydig cell-specific INSL3 were measured as controls. Human gene names are presented. **(B)** Levels of ASPX_HUMAN protein (ACRV1 gene) decreased below the SRM assay LOD (dashed line) in matched post-vasectomy SP (PostV; N=18), as compared to pre-vasectomy SP (PreV; N=18) and suggested the high testis specificity of ASPX_HUMAN. **(C)** ASPX_HUMAN levels measured in the independent SP sets of unmatched pre-vasectomy (N=18), unmatched post-vasectomy (N=18), OA (N=19), as well as NOA samples with the histological subtypes of hypospermatogenesis (HGS; N=5), maturation arrest (MA; N=21) and Sertoli cell-only (SCO; N=12). Interestingly, ASPX_HUMAN was detected in some HSG and MA samples. **(D)** ASPX_HUMAN levels measured in the independent SP set of NOA patients (N=81) with the confirmed mTESE outcomes. Interestingly, ASPX_HUMAN was detected in some mTESE spermatozoa (Spz)- and elongated spermatid (eST)-positive SP samples. Performance of other germ cell-specific proteins is presented in **figs. S1-3**. e+rST, elongated+round spermatids; rST, round spermatids; GC, germ cells.

### Prediction of mTESE outcomes based on the SP levels of 18 proteins

We then measured 18 proteins in SP of 91 NOA patients with the known mTESE outcomes (**fig. S3**). Interestingly, SRM data revealed that ASPX_HUMAN (**Fig. 2D**), TX101_HUMAN, ADA18_HUMAN and ADA30_HUMAN were the best predictors for extraction of spermatozoa or elongated spermatids (32%, 32%, 19% and 18% diagnostic sensitivity, respectively). In addition, ADA18_HUMAN and ADA30_HUMAN levels were independent from TX101_HUMAN and increased the combined diagnostic sensitivity from 32% to 53%, while other proteins revealed no complementarity (**fig. S3**). Even though a multiplex SRM assay could be a viable approach to measure the panel of these soluble SP markers, their relatively low diagnostic sensitivity motivated us to evaluate alternative approaches.

### Selection of organelle-specific proteins

We previously evaluated expression and localization of testis-specific proteins TX101_HUMAN, DPEP3_HUMAN and LY6K_HUMAN in mature spermatozoa *(30, 42)* and revealed their cell surface localization within the post-acrosomal region. In this study, we selected two additional proteins ADA20_HUMAN (ADAM20 gene) and ADA29_HUMAN (ADAM29 gene) due to their elevated expression at the late stage germ cells (**table S2**), their similarity to the sperm-egg binding fertilin beta ADAM2 *(43)*, and availability of suitable antibodies. Immunofluorescent microscopy revealed cell surface localization of ADA20_HUMAN and ADA29_HUMAN within the post-acrosomal region of spermatozoa (**fig. S4**). Finally, we realized that proteins localized on the cell surface within the post-acrosomal region could hardly distinguish spermatozoa heads from the round spermatids within the debris-laden NOA cell pellets or complex mixtures of germ cells and damaged spermatozoa. As a result, we proposed that the ideal markers to identify morphologically intact mature spermatozoa would include proteins found exclusively within the unique organelles of mature spermatozoa, such as acrosomes and tails.

While our candidate markers included some potential acrosome-localized proteins, such as ASPX_HUMAN, there was no indication that any of the 45 proteins (**table S2**) could be localized within the spermatozoa tails. Thus, we re-searched our initial list of testis-specific proteins (**table S1**), the Human Protein Atlas and shotgun mass spectrometry data in order to consider some intracellular proteins. Interestingly, a testis- and late spermatid-specific intracellular protein AKAP4_HUMAN (A-kinase anchoring protein 4) emerged as one of the most abundant human spermatozoa proteins. AKAP4 has previously been identified as a tail-localized protein in mice *(44)*.

As a result, we suggested that simultaneous detection of spermatozoa acrosomes with ASPX_HUMAN, nuclei with the DNA-staining Hoechst dye, and tails with AKAP4_HUMAN could reveal morphologically intact and mature spermatozoa within the highly heterogenous and debris-laden samples of homogenized testicular tissues and semen pellets of NOA patients. Likewise, identification of triple-positive AKAP4^+^/ASPX^+^/Hoechst^+^ cells of elongated shapes would distinguish mature spermatozoa from the early germ cells and round spermatids.

### Subcellular localization of AKAP4_HUMAN and ASPX_HUMAN proteins in spermatozoa

To develop multiplex immunofluorescence microscopy assays, we selected anti-ASPX_HUMAN mouse monoclonal and anti-AKAP4_HUMAN rabbit polyclonal antibodies, as well as secondary antibodies labeled with Alexa 594 and 488, respectively. We confirmed localization of ASPX_HUMAN within acrosomes and ASPX_HUMAN within the tails of the motile normozoospermic spermatozoa (**Figs. 3A-C**), as well as spermatozoa found within the homogenized testicular tissues (**Figs. 3D-F**).

**Fig. 3.**
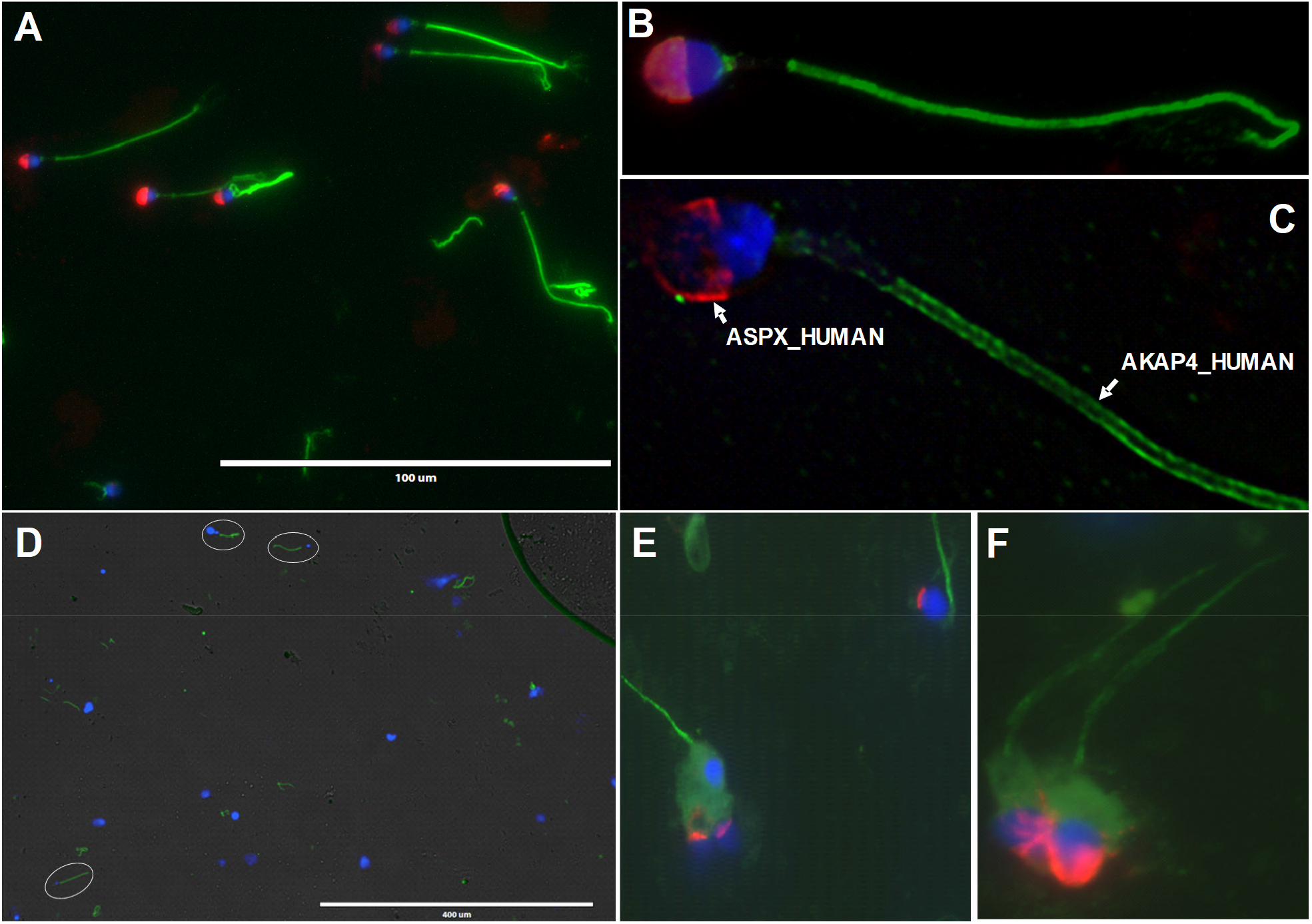
Immunofluorescent microscopy analysis of ASPX_HUMAN and AKAP4_HUMAN proteins in motile spermatozoa and testicular tissues. Fluorescence imaging of AKAP4_HUMAN (green), ASPX_HUMAN (red) and cell nucleus (blue) in motile normozoospermic spermatozoa at 400× (**A**) and 1,000× magnification (**B**). AKAP4_HUMAN and ASPX_HUMAN protein localization in motile spermatozoa visualized and deconvoluted with Huygens software (**C**). Testicular spermatozoa visualized in orchiectomy testicular tissue homogenate at 100× (**D**) and 1,000× (**E, F**) magnification.

### Flow cytometry analysis of AKAP4_HUMAN and ASPX_HUMAN in spermatozoa

Following immunofluorescence microscopy analysis, we optimized ASPX_HUMAN and AKAP4_HUMAN staining for the flow cytometry and imaging flow cytometry analysis. We discovered that flow cytometry analysis required spermatozoa permeabilization with 70% ethanol. Flow cytometry (**Fig. 4**) and imaging flow cytometry (**Fig. 5** and **fig. S5**) analysis revealed simultaneous presence of the high levels of ASPX_HUMAN and AKAP4_HUMAN proteins in >70% of motile normozoospermic spermatozoa.

**Fig. 4.**
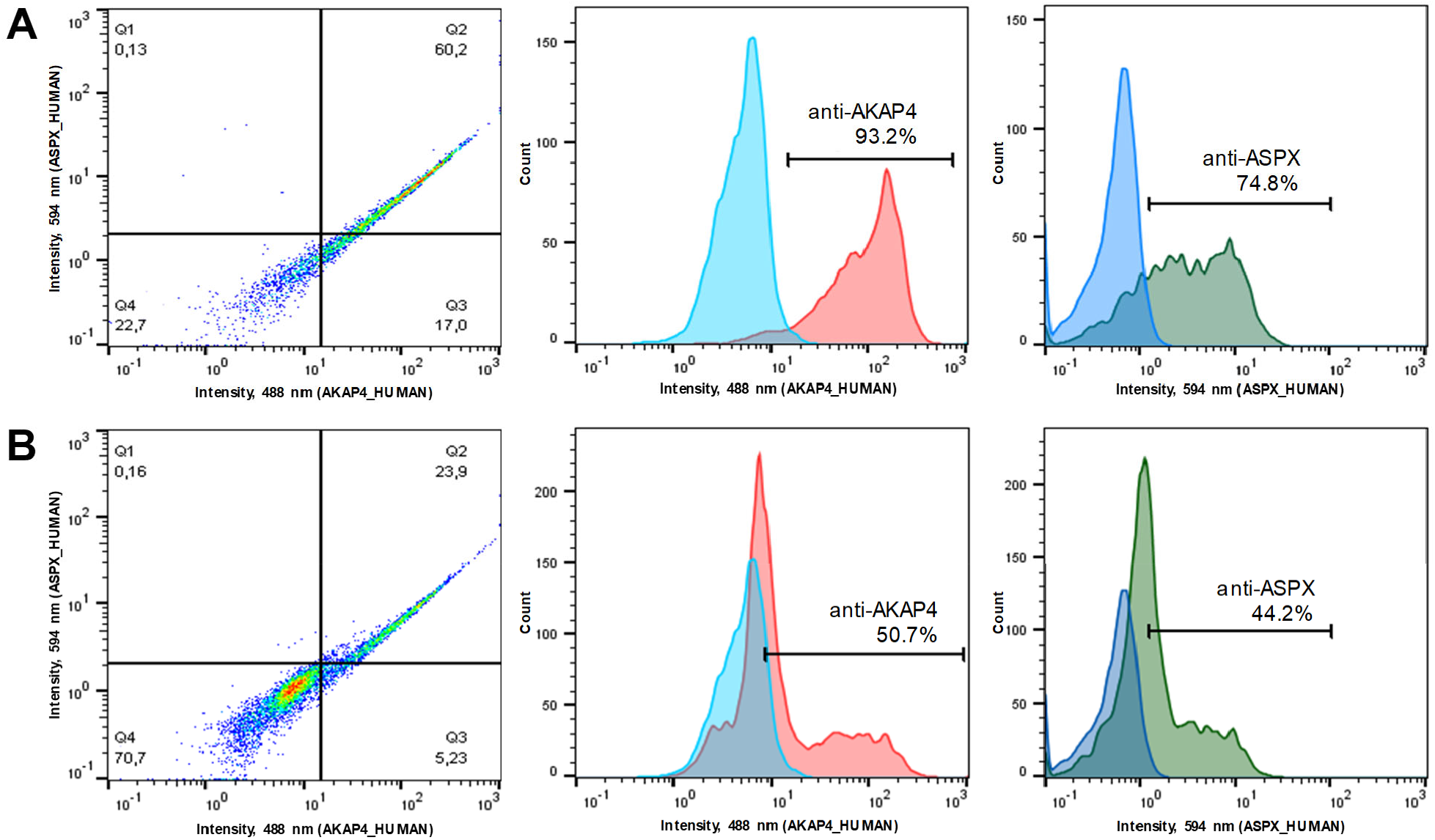
Flow cytometry analysis of AKAP4^+^ and ASPX^+^ spermatozoa. Normozoospermic whole semen pellets were washed, and spermatozoa were treated with 70% ethanol (**A**) or PBS only (**B**) prior to staining with anti-AKAP4 and anti-ASPX primary antibodies and the corresponding secondary antibodies. Ethanol permeabilization significantly increased the fraction of AKAP4^+^/ASPX^+^ spermatozoa.

**Fig. 5.**
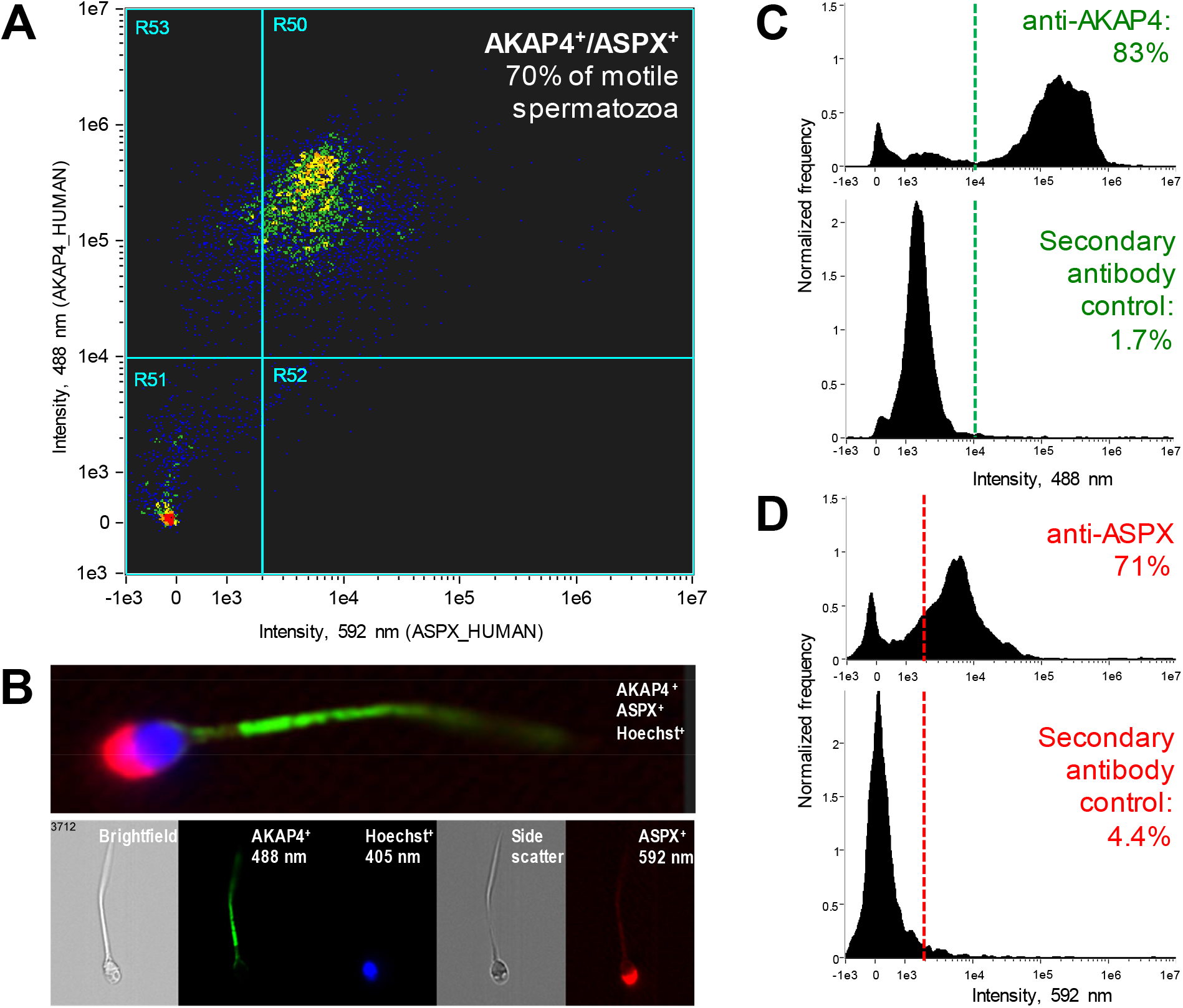
Imaging flow cytometry analysis of AKAP4^+^/ASPX^+^/Hoechst^+^ spermatozoa. (**A**) 70% of motile spermatozoa permeabilized with ethanol were AKAP4^+^/ASPX^+^ positive. (**B**) Fluorescence imaging of motile spermatozoa with AKAP4 (green; Alexa 488), ASPX (red; Alexa 594) and nuclear (blue; Hoechst) staining. (**C, D**) Staining for AKAP4 (83% positive) and ASPX (71% positive) in motile spermatozoa versus secondary antibody-only controls.

### Identification of rare intact AKAP4^+^/ASPX^+^/Hoechst^+^ spermatozoa in NOA semen pellets

Semen pellets of 30 patients (**Table 2**) were subjected to the exhaustive imaging flow cytometry analysis for AKAP4^+^/ASPX^+^/Hoechst^+^ cells of elongated shapes. As a result, numerous AKAP4^+^/ASPX^+^/Hoechst^+^ spermatozoa were identified in semen pellets of normozoospermic and oligozoospermic samples (**fig. S5, 6**). In addition, we identified several AKAP4^+^/ASPX^+^/Hoechst^+^ triple-positive morphologically intact spermatozoa in semen pellets of NOA patients with no spermatozoa identified at the routine laboratory semen analysis (**Fig. 6** and **fig. S7**). Our NOA-mTESE reference set of 7 patients with the known mTESE outcomes provided 83% diagnostic sensitivity at presumed 100% specificity to non-invasively detect intact spermatozoa in the semen pellet of NOA patients. Finally, we evaluated the performance of AKAP4/ASPX/Hoechst imaging flow cytometry with an independent set of azoospermia samples with unknown mTESE outcomes and negative laboratory semen analysis findings. As a result, we identified numerous morphologically intact spermatozoa in the semen pellet of three patients (**Table 2**). Collectively, these results revealed that our multi-step gating strategy and imaging flow cytometry visualization of AKAP4^+^/ASPX^+^/Hoechst^+^ cells bearing elongated tails and acrosome-capped nuclei could enable fast and unambiguous identification of rare and morphologically intact spermatozoa in semen pellets of NOA patients. Pending further validation, our assay may emerge as a non-invasive test to predict the retrieval of morphologically intact spermatozoa by mTESE, thus improving diagnostics and treatment of the severe forms of male infertility.

**Table 2.** Imaging flow cytometry identification of morphologically intact AKAP4^+^/ASPX^+^/Hoechst^+^ spermatozoa in NOA semen pellets. The reference NOA-mTESE set provided 83% sensitivity to non-invasively detect spermatozoa in NOA patients. *unknown NOA or OA subtype.

| **Reference NOA-mTESE set (Murray Koffler Urologic Wellness Centre)** |  |  |  |
| --- | --- | --- | --- |
| **Patient ID** | **mTESE outcome** | **TX101_HUMAN in SP, ng/ml (29)** | **AKAP4<sup>+</sup>/ASPX<sup>+</sup>/Hoechst<sup>+</sup> count** |
| Patient #88 | Spermatozoa | 0.5 | 11 spermatozoa |
| Patient #117 | Spermatozoa | 14.2 | 2 spermatozoa |
| Patient #55 | Spermatozoa | 0.5 | 1 spermatozoon |
| Patient #85 | Spermatozoa | < 0.2 | 1 spermatozoon |
| Patient #93 | Spermatozoa | 504 | 1 spermatozoon |
| Patient #60 | Spermatozoa | < 0.2 | No spermatozoa |
| Patient #29 | No spermatozoa | < 0.2 | No spermatozoa |
| **Additional NOA set (CReATe Fertility Centre)** |  |  |  |
| **Patient ID** | **Patient group** | **Andrology lab count** | **AKAP4<sup>+</sup>/ASPX<sup>+</sup>/Hoechst<sup>+</sup> count** |
| Patient #1 | NOA | No spermatozoa | 77 spermatozoa |
| Patient #2 | NOA | No spermatozoa | 15 spermatozoa |
| Patient #3 | NOA | No spermatozoa | 10 spermatozoa |
| Patient #4 | NOA | No spermatozoa | 2 spermatozoa |
| Patients #5-13 | NOA | No spermatozoa | No spermatozoa |
| Patients #14-20 | Azoospermia* | No spermatozoa | No spermatozoa |
| Patient #21 | OA | No spermatozoa | No spermatozoa |
| Patient #22 | Oligozoospermia | 1.6 million/mL | >3,000 spermatozoa |
| Patient #23 | Normozoospermia | 15 million/mL | Numerous spermatozoa |

**Fig. 6.**
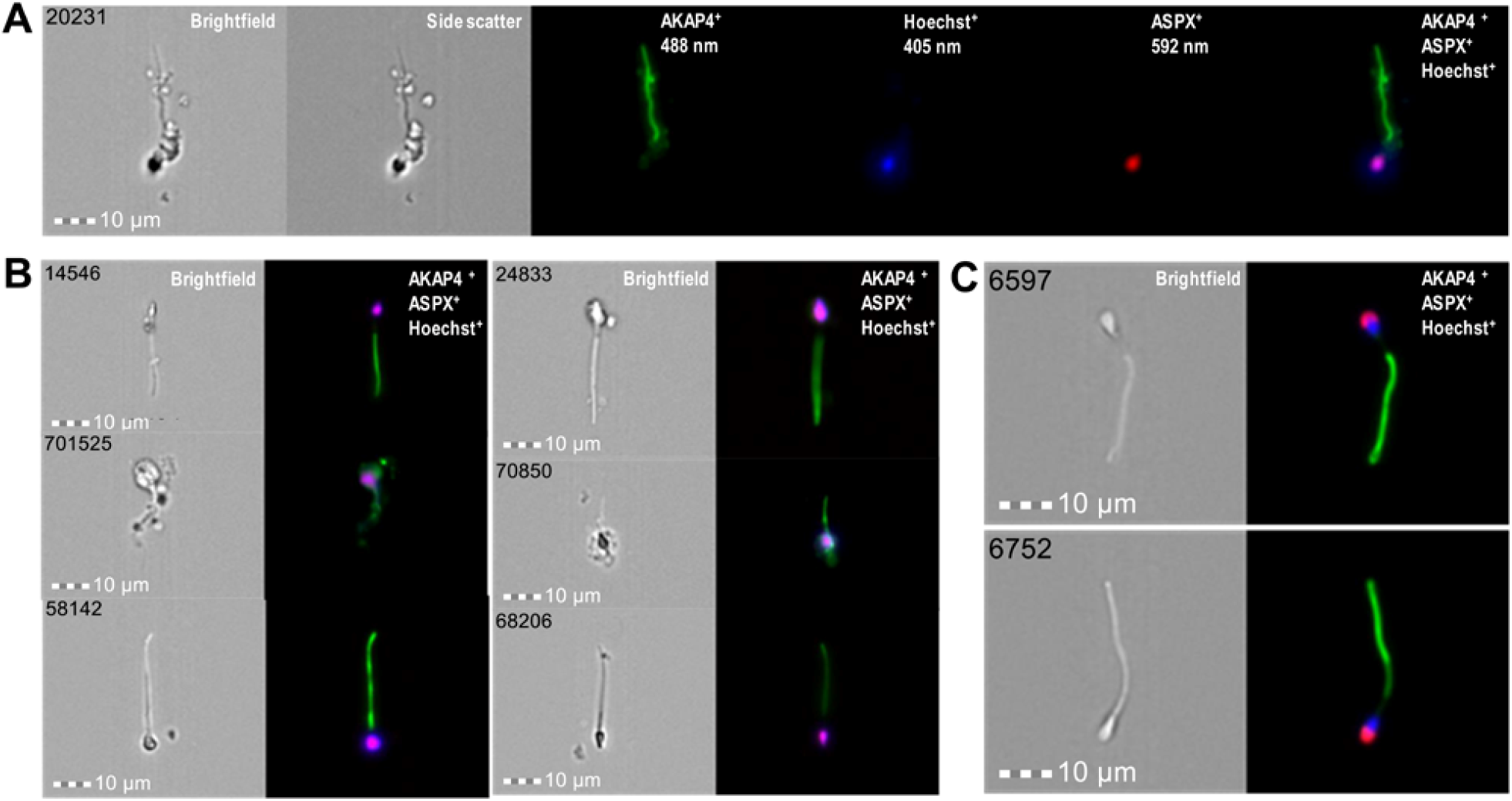
Imaging flow cytometry identification and visualization of morphologically intact AKAP4^+^/ASPX^+^/Hoechst^+^ spermatozoa in semen pellets. (**A, B**) A total of 1,386,670 ImageStream images were recorded during exhaustive 14 runs of a semen pellet sample of NOA patient #88, and 11 morphologically intact AKAP4^+^/ASPX^+^/Hoechst^+^ spermatozoa were identified. (**C**) A single run of 21,375 images of a semen pellet sample of oligozoospermic patient #22 revealed numerous AKAP4^+^/ASPX^+^/Hoechst^+^ spermatozoa. Additional images are presented in **figs. S5-S7**.

## DISCUSSION

In this study, we hypothesized that some testis- and germ cell-specific proteins could emerge as markers for the non-invasive and highly specific identification of rare spermatozoa within the debris-laden semen pellets of NOA patients. Our approach for selection of the most promising testis- and germ cell-specific proteins relied on re-analysis of our previously identified spermatozoa *(30)* and seminal plasma proteomes *(33)*, and on mining of the well-established proteomic databases. Since the high-quality protein assays are essential for biomarker discovery and development *(45-47)*, we suggested that targeted proteomic assays due to their rapid design, development and execution could be particularly valuable tools to evaluate clinical utility of proteins lacking high-quality immunoassays or antibodies. We previously demonstrated that targeted proteomic assays provided robust quantification of proteins in human cell lines *(48, 49)*, primary cells *(50, 51)*, tissues *(38)*, biological fluids *(52-56)*, and serum *(57-59)*. Likewise, quantitative proteomic assays to evaluate testis-specific proteins in spermatozoa, vas deferens fluid and seminal plasma would enable experimental verification and rapid prioritization of candidates. Immunofluorescence microscopy and flow cytometry assays would independently confirm subcellular localization and expression levels of the most promising candidates.

Recent-omics studies reported that nearly 6% of all human proteins were expressed in testis *(34)*. Unique and highly specialized roles of testis-specific proteins in spermatogenesis, remodeling of spermatozoa cell surface, sperm transit, sperm-oocyte interaction and fusion were discovered *(60)*. Prominent examples included metalloprotease-disintegrin ADAM2 *(61)*, the cell adhesion tetraspanin CD9 *(62)* and the sperm-egg fusion protein IZUMO1 *(17)*. Discovery of the cell surface recognition complex of IZUMO1 protein and the sperm-egg fusion protein JUNO provided detailed insights into gamete recognition and sperm-oocyte fusion *(63, 64)*.

We previously identified and validated numerous testis-specific proteins as biomarkers for the differential diagnosis of azoospermia *(25, 26)*. The combination of a testis- and germ cell-specific protein TX101_HUMAN and an epididymis-specific protein ECM1_HUMAN provided 81% sensitivity at 100% specificity to differentiate between non-obstructive and obstructive azoospermia *(28)*. However, an initial study *(28)* revealed only moderate performance of TX101_HUMAN to predict spermatozoa retrieval by mTESE (73% sensitivity and 64% specificity at the assay limit of detection ≥0.6 ng/mL). A follow-up study *(29)* with an improved immunoassay revealed 87% sensitivity and 53% specificity at ≥0.2 ng/mL to extract mature spermatozoa.

Our success with azoospermia biomarkers motivated us to extend our search for additional testis- and germ cell-specific proteins and to identify more specific and sensitive biomarkers for prediction of mTESE outcomes. Integration of our experimental proteomic data with mining of the Human Protein Atlas *(34)* and NextProt *(35)* data revealed that spermatozoa expressed 578 testis-specific proteins, of which 129 and 55 proteins had at least a single membrane-bound or secreted isoform, respectively. Our shotgun mass spectrometry data on protein expression and tryptic peptide fragmentation enabled prioritization of candidate proteins and selection of proteotypic peptides suitable for development of targeted proteomic assays.

Experimental verification excluded poor markers, confirmed our previous findings for TX101_HUMAN and DPEP3_HUMAN, and provided ASPX_HUMAN as an additional marker with AUC=1.0 to detect post-vasectomy or OA. ASPX_HUMAN emerged as the most promising marker for the subsequent validation due to: (i) late germ cell-specificity and subcellular localization in the acrosomal versus post-acrosomal regions of spermatozoa; (i) detection in NOA-maturation arrest (**Fig. 2C**) and mTESE spermatozoa/elongated spermatid-positive SP samples (**Fig. 2D**); (iii) availability of high-quality monoclonal antibodies to detect the native non-denaturated protein forms. Likewise, presumably intracellular AKAP4_HUMAN protein could be readily detected by immunofluorescence microscopy and flow cytometry within the spermatozoa tails.

ASPX_HUMAN and AKAP4_HUMAN proteins emerged as the best combination for the highly specific detection and visualization of the morphologically intact spermatozoa by the multiplex imaging flow cytometry. Analysis of normozoospermic and oligozoospermic samples (**figs. S5-6)** facilitated development of a multi-step gating strategy based on spermatozoa brightfield area and aspect ratio, AKAP4 and Hoechst fluorescence intensity, AKAP4 aspect ratio intensity sorting, and rapid visual inspection of the combined AKAP4^+^/ASPX^+^/Hoechst^+^ images. The combined approach facilitated analysis of millions of single cell images, straightforward visualization and rapid identification of the rare morphologically intact triple-positive AKAP4^+^/ASPX^+^/Hoechst^+^ spermatozoa (as few as 1 spermatozoa in ∼100,000 ImageStream images) even within the debris-laden NOA semen pellets (**fig. S6**).

It should be emphasized that previous studies on the soluble mTESE biomarkers in SP, including our studies *(28, 29)*, revealed relatively high diagnostic sensitivity, but suffered from the low diagnostic specificity to predict spermatozoa retrieval. Interestingly, low diagnostic specificity was found even for the highly testis-specific proteins measured in SP *(28)*, likely due to their presence at the continuum of germ cell differentiation, including early and late germ cells (**Table 1**). Unlike measurements of soluble markers in SP, AKAP4^+^/ASPX^+^/Hoechst^+^ imaging flow cytometry provided additional visual information for the unique morphology of spermatozoa including acrosome-capped nuclei and elongated tails. Manual inspection of few AKAP4^+^/ASPX^+^/Hoechst^+^ cells provided nearly unambiguous identification of mature spermatozoa and resulted in nearly no false-positives and high diagnostic specificity unachievable in the previous studies.

In future, combination of our less expensive TX101_HUMAN immunoassay in SP as an initial screening approach with 87% sensitivity *(28, 29)* and the more expensive AKAP4^+^/ASPX^+^/Hoechst^+^ assay with high specificity and positive predictive values to follow up on TX101_HUMAN positive result could emerge as a viable clinical strategy for non-invasive prediction of mTESE outcomes. However, an independent prospective validation of such combination and corresponding diagnostic cutoffs in large sets of samples will be required. As potential limitations, it should be noted that rare AKAP4^+^/ASPX^+^/Hoechst^+^ spermatozoa selected by imaging flow sorting approaches may not be suitable for ICSI due to the requirement for ethanol permeabilization and the residual anti-AKAP4 and ASPX antibodies bound to spermatozoa.

Our work may represent one of the most comprehensive studies on testis- and germ cell-specific proteins. Since no immunoassays are currently available for most germ cell-specific proteins evaluated in our study, our SRM assays are first-of-its-kind analytical tools to measure these germ cell-specific proteins in semen, SP, vas deferens fluid and spermatozoa. Some of these proteins could be essential for the spermatozoa-oocyte interaction *(65)* and emerge as targets to develop non-hormonal male contraceptives *(66)*, or emerge as non-invasive compound biomarkers to evaluate male contraceptives. The reported panel of proteins will facilitate evaluation of spermatozoa and acrosome integrity and enable future functional and translational studies on the biology of human reproduction, spermatogenesis, and male fertility.

## MATERIALS AND METHODS

### Study design

The objectives of this study were to (i) identify testis-, germ cell- and organelle-specific proteins; (ii) retrospectively evaluate expression of candidate markers by targeted proteomic assays, immunofluorescence microscopy and imaging flow cytometry in semen of normozoospermic and NOA patients; (iii) develop a multiplex imaging flow cytometry assay for non-invasive identification of rare spermatozoa in semen of NOA patients. According to power calculations (**table S6**), 80% power to detect highly testis-specific proteins, such as TX101_HUMAN *(25)*, can be achieved with 16 independent pre-vasectomy versus 16 post-vasectomy, or OA, or NOA samples (α=0.05, two-tailed t-test; TX101_HUMAN levels and standard deviations from our previous study *(25)*). To be able to establish sensitivities and specificities of each protein with higher accuracy, we evaluated our panel of testis-specific proteins in larger sets of SP samples already available in our lab (36 pre-vasectomy, 36 pos-vasectomy, 19 OA, 38 NOA with histological subtypes, and 91 NOA with the known mTESE outcome).

### Patients

Semen, spermatozoa, SP and orchiectomy testicular tissues (**table S7**) with relevant clinical information were collected with institutional REB-approved consent at Murray Koffler Urologic Wellness Centre at Sinai Health System (REB# 15-0149-A) and CReATe Fertility Centre (University of Toronto REB# 09-0156-E30252 and Sinai Health System REB# 14-0032-E). Men with normal spermatogenesis were confirmed fertile men with normal sperm count by semen analysis (>15 million per mL according to the World Health Organization reference values). These men were referred for vasectomy, and SP samples were obtained prior to vasectomy. Post-vasectomy group included SP samples obtained from fertile men 3 to 6 months after vasectomy, and zero sperm count was confirmed by at least two semen analyses. OA group included men with biopsy-confirmed OA, obstruction at the epididymis, normal testicular volume and normal FSH (1-18 IU/L). The NOA group included men with azoospermia by semen analysis and elevated FSH (>18 IU/L) or NOA confirmed by testicular biopsy. Y chromosome deletion status was not used as an independent parameter for NOA cases. Patients were included if they had medical indications for the mTESE and IVF/ICSI procedures and had no contra-indications. Female infertility factors were excluded.

### Clinical samples

Semen samples from men with normal spermatogenesis pre- and post-vasectomy and men with azoospermia were collected after a minimum of 3 days of sexual abstinence. The sample collection procedure was identical for all sets. For proteomics measurements, semen was left to liquefy at room temperature for 1 hour, then aliquoted in 1 mL portions, and centrifuged once at 800 g to pellet spermatozoa, and then centrifuged twice at 13,000 g to remove cellular debris and obtain SP devoid of cells and cellular components. Spermatozoa pellet and SP were stored at −80 °C until use. To obtain motile spermatozoa, semen was incubated at 37ºC for 15 min, followed by one-layer density-gradient centrifugation. The spermatozoa pellet was washed twice with PBS and either cryopreserved or fixed for immunofluorescence microscopy and flow cytometry. The samples were anonymous and labeled with biobank identification numbers. The research laboratory did not have any identifying patient information but was provided with the results of the regular semen analysis. Testicular sperm was isolated from testicular tissues following mechanical dissociation and digestion by collagenase (1.4 mg/mL; #GM501, Gynemed) diluted with HTF HEPES media (#2002, InVitroCare, Frederick, MD).

### SRM targeted proteomic assays

Semen, spermatozoa, and SP samples were lysed with Rapigest SF surfactant at 60 °C *(30, 48)*. Proteins were denatured, reduced, alkylated, and digested by trypsin. Tryptic peptides (∼10 μg) were separated by reverse-phase chromatography and quantified by SRM targeted proteomic assays. SRM assays were developed for TSQ Quantiva mass spectrometer (Thermo Scientific) as previously reported *(52-55, 67-70)*. The best proteotypic peptides were selected based on their relative intensities in spermatozoa and SP proteomes. Peptide sequence specificity was confirmed with the Protein BLAST. Heavy isotope-labeled peptide internal standards were used for assay development, selection of the best precursor-to-fragment ion transitions, and for the accurate relative quantification. A single analytical (full process digestion and sample preparation) and two technical replicates were analyzed by LC-SRM for each SP sample. Single analytical replicates were justified by the established proteomic protocols for SP samples and by the expected dramatic decrease of testis-specific protein concentrations in post-vasectomy, OA and NOA samples, as compared to pre-vasectomy samples *(25)*. SRM area was used to calculate L/H ratios, protein concentrations and standard deviations. To limit the effects of LC carry-over (∼0.2%), a restricted randomized block design was used for randomization. SP samples were split into pre-vasectomy, post-vasectomy, OA, or NOA blocks of 4×2 samples, and blocks were randomized on 96-well plates for sample preparation and LC-SRM analysis.

### Immunofluorescence

Approximately 100 spermatozoa per slide were fixed with 4% paraformaldehyde, permeabilized with 70% ethanol, and mounted onto microscope slides for immunofluorescence staining. Mouse monoclonal antibodies included: anti-ACRV1 (1 μg/mL final; Sinobiological; 11789-MM01), anti-ADAM20 (10 μg/mL final; Abnova; H00008748-M05) and anti-ADAM29 (10 μg/mL final; Abnova; H00011086-M09). Rabbit polyclonal and the corresponding secondary antibodies included anti-AKAP4 (5 μg/mL final; Thermo Fisher; PA5-38015), Alexa Fluor 594 goat-anti-mouse (2 μg/mL final; Thermo Fisher; A-21125) and Alexa Fluor 488 goat-anti-rabbit (1 μg/mL final; Thermo Fisher; A-11034). High-magnification fluorescence microscopy (100×; Olympus BX61) was used to visualize protein markers and DAPI staining of nuclei (Thermo Fisher; S36968), as previously described *(30)*.

### Flow cytometry

Whole semen and washed motile spermatozoa were analyzed for ASPX_HUMAN and AKAP4_HUMAN expression using flow cytometry (MACSQuant Analyzer 10; Miltenyi Biotec, Germany). Data analysis was performed using FlowJo™ v10 software (BD Life Sciences).

### Imaging flow cytometry

Imaging flow cytometry (Amnis ImageStreamX Mark II, Luminex) was used to evaluate the performance of tail- and acrosome-specific proteins and detect rare spermatozoa in semen pellets of azoospermic and oligozoospermic patients. Semen cell pellets were washed with PBS and stained with Hoechst 33342 and primary antibodies (mouse anti-ASPX_HUMAN #11789-MM01 and rabbit anti-AKAP4 #PA5-38015), followed by the secondary antibodies labeled with Alexa Fluor 594 and 488, respectively. ImageStream imaging flow cytometry (brightfield and sidescatter imaging, and fluorescence excitation with 405, 488 and 592 nm lasers) visualized cells (up to 100,000 images per run; up to 14 runs per NOA semen pellets). Image analysis was performed with IDEAS v6.2 software (Luminex). Analysis of normozoospermic and oligozoospermic samples facilitated development of a sequential gating template based on the brightfield area and aspect ratio, AKAP4 and Hoechst fluorescence intensity, followed by AKAP4 aspect ratio intensity sorting and visual inspection of the combined AKAP4^+^/ASPX^+^/Hoechst^+^ images. Such multi-step gating strategy facilitated manual inspection of thousands of images and rapid identification of the morphologically intact AKAP4^+^/ASPX^+^/Hoechst^+^ spermatozoa.

### Statistical analysis

GraphPad PRISM v5.03 was used to calculate ROC AUC area, sensitivity and specificity. A one-tailed Mann-Whitney U test was used for pairwise comparisons between groups, and P-values <0.05 were considered significant. G*Power software (version 3.1.9.6, Heinrich Heine University Dusseldorf) enabled power calculations. Presented study was based on the retrospectively collected semen and SP samples, and followed the Standards for Reporting Diagnostic Accuracy Studies (STARD 2015) recommendations (**table S8**).

## Data Availability

All data produced in the present study are available upon reasonable request to the authors

## List of Supplementary Materials

Fig. S1. SRM analysis of the matched pre-vasectomy (N=18) and post-vasectomy (N=18) SP to access testis/epididymis specificity of candidate proteins.

Fig. S2. SRM quantification of candidate proteins in SP of pre-vasectomy, post-vasectomy and NOA patients.

Fig. S3. SRM quantification of candidate proteins in SP of NOA patients with the known mTESE outcomes.

Fig. S4. Localization of ADA20_HUMAN and ADA29_HUMAN proteins in motile spermatozoa.

Fig. S5. Imaging flow cytometry identification and visualization of the morphologically normal and intact AKAP4^+^/ASPX^+^/Hoechst^+^ spermatozoa in semen pellet of a normozoospermic patient.

Fig. S6. Imaging flow cytometry identification and visualization of intact AKAP4^+^/ASPX^+^/Hoechst^+^ spermatozoa in semen pellet of a patient diagnosed with oligospermia.

Fig. S7. Visualization of two intact AKAP4^+^/ASPX^+^/Hoechst^+^ spermatozoa in NOA semen pellet.

Table S1. Selected 578 testis-specific proteins and their relative abundance in human spermatozoa.

Table S2. Selected 45 testis germ cell-specific proteins expressed at the different stages of spermatogenesis.

Table S3. Heavy isotope-labeled tryptic peptide internal standards used for SRM assay development.

Table S4. SRM assays for quantification of 45 testis germ cell-specific proteins.

Table S5. SRM assays for quantification of 18 testis germ cell-specific proteins.

Table S6. Power calculations.

Table S7. Summary of clinical samples analyzed in the present study.

Table S8. STARD 2015 recommendations.

## Acknowledgements

We thank Susan Lau (Sinai Health System) and CReATe andrology lab personnel for assistance with the clinical samples. We thank the Advanced Center for Detection of Cancer mass spectrometry facility (Sinai Health System) and Lunenfeld-Tanenbaum Research Institute Flow Cytometry facility for the cost-recovery mass spectrometry and imaging flow cytometry services.

## Author contributions

A.P.D. designed the research project; A.P.D., J.Z. and M.K. performed the experiments; A.P.D. and Z.F. analyzed data; K.J., S.M. and C.L. provided samples and clinical information; A.P.D. wrote manuscript, and all authors contributed to the revisions.

## Data Availability

Raw SRM data, as well as processed Skyline files, were deposited to Peptide Atlas with the dataset identifier PASS01755 (www.peptideatlas.org/PASS/PASS01755).

## Non-standard abbreviations

AUC: area under the receiver operating characteristic curve
ICSI: intracytoplasmic sperm injection
IVF: in vitro fertilization
LC-MS/MS: liquid chromatography - tandem mass spectrometry
LOD: limit of detection
mTESE: microdissection testicular sperm extraction
MWU: Mann Whitney Unpaired t-test
NOA: non-obstructive azoospermia
NPV: negative predictive value
PPV: positive predictive value
SP: seminal plasma
SRM: selected reaction monitoring

## References

1. Report on evaluation of azoospermic male. Fertil. Steril. 86, S210–S215 (2006).

2. WHO Laboratory Manual for the Examination of Human Semen and Sperm-Cervical Mucus Interaction. (Cambridge University Press, ed. 4, 1999), pp. 106.

3. S. von Eckardstein, M. Simoni, M. Bergmann, G. F. Weinbauer, P. Gassner, A. G. Schepers, E. Nieschlag, Serum inhibin B in combination with serum follicle-stimulating hormone (FSH) is a more sensitive marker than serum FSH alone for impaired spermatogenesis in men, but cannot predict the presence of sperm in testicular tissue samples. J Clin Endocrinol Metab 84, 2496–2501 (1999).

4. K. Jarvi, K. Lo, A. Fischer, J. Grantmyre, A. Zini, V. Chow, V. Mak, CUA Guideline: The workup of azoospermic males. Can Urol Assoc J 4, 163–167 (2010).

5. P. N. Schlegel, G. D. Palermo, M. Goldstein, S. Menendez, N. Zaninovic, L. L. Veeck, Z. Rosenwaks, Testicular sperm extraction with intracytoplasmic sperm injection for nonobstructive azoospermia. Urology 49, 435–440 (1997).

6. P. N. Schlegel, Testicular sperm extraction: microdissection improves sperm yield with minimal tissue excision. Hum Reprod 14, 131–135 (1999).

7. R. Ramasamy, P. N. Schlegel, Microdissection testicular sperm extraction: effect of prior biopsy on success of sperm retrieval. J Urol 177, 1447–1449 (2007).

8. P. N. Schlegel, Causes of azoospermia and their management. Reprod Fertil Dev 16, 561–572 (2004).

9. C. Kang, N. Punjani, P. N. Schlegel, Reproductive Chances of Men with Azoospermia Due to Spermatogenic Dysfunction. J Clin Med 10, (2021).

10. A. N. Yatsenko, A. P. Georgiadis, A. Ropke, A. J. Berman, T. Jaffe, M. Olszewska, B. Westernstroer, J. Sanfilippo, M. Kurpisz, A. Rajkovic, S. A. Yatsenko, S. Kliesch, S. Schlatt, F. Tuttelmann, X-linked TEX11 mutations, meiotic arrest, and azoospermia in infertile men. N Engl J Med 372, 2097–2107 (2015).

11. Y. Shen, F. Zhang, F. Li, X. Jiang, Y. Yang, X. Li, W. Li, X. Wang, J. Cheng, M. Liu, X. Zhang, G. Yuan, X. Pei, K. Cai, F. Hu, J. Sun, L. Yan, L. Tang, C. Jiang, W. Tu, J. Xu, H. Wu, W. Kong, S. Li, K. Wang, K. Sheng, X. Zhao, H. Yue, X. Yang, W. Xu, Loss-of-function mutations in QRICH2 cause male infertility with multiple morphological abnormalities of the sperm flagella. Nat Commun 10, 433 (2019).

12. C. Coutton, A. S. Vargas, A. Amiri-Yekta, Z. E. Kherraf, S. F. Ben Mustapha, P. Le Tanno, C. Wambergue-Legrand, T. Karaouzene, G. Martinez, S. Crouzy, A. Daneshipour, S. H. Hosseini, V. Mitchell, L. Halouani, O. Marrakchi, M. Makni, H. Latrous, M. Kharouf, J. F. Deleuze, A. Boland, S. Hennebicq, V. Satre, P. S. Jouk, N. Thierry-Mieg, B. Conne, D. Dacheux, N. Landrein, A. Schmitt, L. Stouvenel, P. Lores, E. El Khouri, S. P. Bottari, J. Faure, J. P. Wolf, K. Pernet-Gallay, J. Escoffier, H. Gourabi, D. R. Robinson, S. Nef, E. Dulioust, R. Zouari, M. Bonhivers, A. Toure, C. Arnoult, P. F. Ray, Mutations in CFAP43 and CFAP44 cause male infertility and flagellum defects in Trypanosoma and human. Nat Commun 9, 686 (2018).

13. K. Siklenka, S. Erkek, M. Godmann, R. Lambrot, S. McGraw, C. Lafleur, T. Cohen, J. Xia, M. Suderman, M. Hallett, J. Trasler, A. H. Peters, S. Kimmins, Disruption of histone methylation in developing sperm impairs offspring health transgenerationally. Science 350, aab2006 (2015).

14. M. Jodar, E. Sendler, S. I. Moskovtsev, C. L. Librach, R. Goodrich, S. Swanson, R. Hauser, M. P. Diamond, S. A. Krawetz, Absence of sperm RNA elements correlates with idiopathic male infertility. Sci Transl Med 7, 295re296 (2015).

15. J. Zhang, X. Mu, Y. Xia, F. L. Martin, W. Hang, L. Liu, M. Tian, Q. Huang, H. Shen, Metabolomic analysis reveals a unique urinary pattern in normozoospermic infertile men. J Proteome Res 13, 3088–3099 (2014).

16. W. Yu, H. Zheng, W. Lin, A. Tajima, Y. Zhang, X. Zhang, H. Zhang, J. Wu, D. Han, N. A. Rahman, K. S. Korach, G. F. Gao, I. Inoue, X. Li, Estrogen promotes Leydig cell engulfment by macrophages in male infertility. J Clin Invest 124, 2709–2721 (2014).

17. N. Inoue, M. Ikawa, A. Isotani, M. Okabe, The immunoglobulin superfamily protein Izumo is required for sperm to fuse with eggs. Nature 434, 234–238 (2005).

18. L. Hetherington, E. K. Schneider, C. Scott, D. DeKretser, C. H. Muller, H. Hondermarck, T. Velkov, M. A. Baker, Deficiency in Outer Dense Fiber 1 is a marker and potential driver of idiopathic male infertility. Mol Cell Proteomics 15, 3685–3693 (2016).

19. R. Diao, K. L. Fok, H. Chen, M. K. Yu, Y. Duan, C. M. Chung, Z. Li, H. Wu, Z. Li, H. Zhang, Z. Ji, W. Zhen, C. F. Ng, Y. Gui, Z. Cai, H. C. Chan, Deficient human beta-defensin 1 underlies male infertility associated with poor sperm motility and genital tract infection. Sci Transl Med 6, 249ra108 (2014).

20. J. M. Bieniek, A. P. Drabovich, K. C. Lo, Seminal biomarkers for the evaluation of male infertility. Asian J Androl 18, 426–433 (2016).

21. A. P. Drabovich, P. Saraon, K. Jarvi, E. P. Diamandis, Seminal plasma as a diagnostic fluid for male reproductive system disorders. Nat Rev Urol 11, 278–288 (2014).

22. C. G. Schiza, K. Jarv, E. P. Diamandis, A. P. Drabovich, An Emerging Role of TEX101 Protein as a Male Infertility Biomarker. EJIFCC 25, 9–26 (2014).

23. A. Tsujimura, K. Matsumiya, Y. Miyagawa, T. Takao, K. Fujita, M. Koga, M. Takeyama, H. Fujioka, A. Okuyama, Prediction of successful outcome of microdissection testicular sperm extraction in men with idiopathic nonobstructive azoospermia. J Urol 172, 1944–1947 (2004).

24. V. Vernaeve, H. Tournaye, J. Schiettecatte, G. Verheyen, A. Van Steirteghem, P. Devroey, Serum inhibin B cannot predict testicular sperm retrieval in patients with non-obstructive azoospermia. Hum Reprod 17, 971–976 (2002).

25. A. P. Drabovich, A. Dimitromanolakis, P. Saraon, A. Soosaipillai, I. Batruch, B. Mullen, K. Jarvi, E. P. Diamandis, Differential diagnosis of azoospermia with proteomic biomarkers ECM1 and TEX101 quantified in seminal plasma. Sci Transl Med 5, 212ra160 (2013).

26. A. P. Drabovich, K. Jarvi, E. P. Diamandis, Verification of male infertility biomarkers in seminal plasma by multiplex selected reaction monitoring assay. Mol Cell Proteomics 10, M110.004127 (2011).

27. C. Bohring, I. Schroeder-Printzen, W. Weidner, W. Krause, Serum levels of inhibin B and follicle-stimulating hormone may predict successful sperm retrieval in men with azoospermia who are undergoing testicular sperm extraction. Fertil Steril 78, 1195–1198 (2002).

28. D. Korbakis, C. Schiza, D. Brinc, A. Soosaipillai, T. D. Karakosta, C. Legare, R. Sullivan, B. Mullen, K. Jarvi, E. P. Diamandis, A. P. Drabovich, Preclinical evaluation of a TEX101 protein ELISA test for the differential diagnosis of male infertility. BMC Med 15, 60 (2017).

29. S. P. Jarvi KA, Schiza C, Drabovich A, Lau S, Soosaipillai A, Korbakis D, Brinc D, Mullen B, Diamandis E, Semen biomarker TEX101 predicts sperm retrieval success for men with testicular failure. F1000Research 10, 569 (2021).

30. C. Schiza, D. Korbakis, K. Jarvi, E. P. Diamandis, A. P. Drabovich, Identification of TEX101-associated proteins through proteomic measurement of human spermatozoa homozygous for the missense variant rs35033974. Mol Cell Proteomics 18, 338–351 (2019).

31. I. Batruch, I. Lecker, D. Kagedan, C. R. Smith, B. J. Mullen, E. Grober, K. C. Lo, E. P. Diamandis, K. A. Jarvi, Proteomic analysis of seminal plasma from normal volunteers and post-vasectomy patients identifies over 2000 proteins and candidate biomarkers of the urogenital system. J Proteome Res 10, 941–953 (2011).

32. I. Batruch, C. R. Smith, B. J. Mullen, E. Grober, K. C. Lo, E. P. Diamandis, K. A. Jarvi, Analysis of seminal plasma from patients with non-obstructive azoospermia and identification of candidate biomarkers of male infertility. J Proteome Res, Epub Feb 2 (2011).

33. A. P. Drabovich, P. Saraon, M. Drabovich, T. D. Karakosta, A. Dimitromanolakis, M. E. Hyndman, K. Jarvi, E. P. Diamandis, Multi-omics Biomarker Pipeline Reveals Elevated Levels of Protein-glutamine Gamma-glutamyltransferase 4 in Seminal Plasma of Prostate Cancer Patients. Mol Cell Proteomics 18, 1807–1823 (2019).

34. M. Uhlen, L. Fagerberg, B. M. Hallstrom, C. Lindskog, P. Oksvold, A. Mardinoglu, A. Sivertsson, C. Kampf, E. Sjostedt, A. Asplund, I. Olsson, K. Edlund, E. Lundberg, S. Navani, C. A. Szigyarto, J. Odeberg, D. Djureinovic, J. O. Takanen, S. Hober, T. Alm, P. H. Edqvist, H. Berling, H. Tegel, J. Mulder, J. Rockberg, P. Nilsson, J. M. Schwenk, M. Hamsten, K. von Feilitzen, M. Forsberg, L. Persson, F. Johansson, M. Zwahlen, G. von Heijne, J. Nielsen, F. Ponten, Proteomics. Tissue-based map of the human proteome. Science 347, 1260419 (2015).

35. P. Gaudet, P.-A. Michel, M. Zahn-Zabal, A. Britan, I. Cusin, M. Domagalski, P. D. Duek, A. Gateau, A. Gleizes, V. Hinard, The neXtProt knowledgebase on human proteins: 2017 update. Nucleic acids research 45, D177–D182 (2017).

36. J. A. Foster, K. L. Klotz, C. J. Flickinger, T. S. Thomas, R. M. Wright, J. R. Castillo, J. C. Herr, Human SP-10: acrosomal distribution, processing, and fate after the acrosome reaction. Biol Reprod 51, 1222–1231 (1994).

37. E. Bjorling, M. Uhlen, Antibodypedia, a portal for sharing antibody and antigen validation data. Mol Cell Proteomics 7, 2028–2037 (2008).

38. Z. Fu, Y. Rais, T. A. Bismar, M. E. Hyndman, X. C. Le, A. P. Drabovich, Mapping Isoform Abundance and Interactome of the Endogenous TMPRSS2-ERG Fusion Protein by Orthogonal Immunoprecipitation-Mass Spectrometry Assays. Mol Cell Proteomics, 100075 (2021).

39. Y. Rais, Z. Fu, A. P. Drabovich, Mass spectrometry-based proteomics in basic and translational research of SARS-CoV-2 coronavirus and its emerging mutants. Clin Proteomics 18, 19 (2021).

40. P. Saraon, D. Cretu, N. Musrap, G. S. Karagiannis, I. Batruch, A. P. Drabovich, T. van der Kwast, A. Mizokami, C. Morrissey, K. Jarvi, E. P. Diamandis, Quantitative proteomics reveals that enzymes of the ketogenic pathway are associated with prostate cancer progression. Mol Cell Proteomics 12, 1589–1601 (2013).

41. P. Saraon, N. Musrap, D. Cretu, G. S. Karagiannis, I. Batruch, C. Smith, A. P. Drabovich, D. Trudel, T. van der Kwast, C. Morrissey, K. A. Jarvi, E. P. Diamandis, Proteomic profiling of androgen-independent prostate cancer cell lines reveals a role for protein S during the development of high grade and castration-resistant prostate cancer. J Biol Chem 287, 34019–34031 (2012).

42. C. Schiza, D. Korbakis, E. Panteleli, K. Jarvi, A. P. Drabovich, E. P. Diamandis, Discovery of a human testis-specific protein complex TEX101-DPEP3 and selection of its disrupting antibodies. Mol Cell Proteomics 17, 2480–2495 (2018).

43. C. Cho, D. O. Bunch, J. E. Faure, E. H. Goulding, E. M. Eddy, P. Primakoff, D. G. Myles, Fertilization defects in sperm from mice lacking fertilin beta. Science 281, 1857–1859 (1998).

44. K. Miki, W. D. Willis, P. R. Brown, E. H. Goulding, K. D. Fulcher, E. M. Eddy, Targeted disruption of the Akap4 gene causes defects in sperm flagellum and motility. Dev Biol 248, 331–342 (2002).

45. A. P. Drabovich, E. Martinez-Morillo, E. P. Diamandis, Toward an integrated pipeline for protein biomarker development. Biochim Biophys Acta 1854, 677–686 (2015).

46. A. P. Drabovich, M. P. Pavlou, I. Batruch, E. P. Diamandis, in Proteomic and Metabolomic Approaches to Biomarker Discovery, H. J. Issaq, T. D. Veenstra, Eds. (Academic Press (Elsevier), Waltham, MA, 2013), chap. 2, pp. 17–37.

47. A. P. Drabovich, E. Martínez-Morillo, E. P. Diamandis, in Proteomics for Biological Discovery. (2019), pp. 63–88.

48. A. P. Drabovich, M. P. Pavlou, A. Dimitromanolakis, E. P. Diamandis, Quantitative analysis of energy metabolic pathways in MCF-7 breast cancer cells by selected reaction monitoring assay. Mol Cell Proteomics 11, 422–434 (2012).

49. A. P. Drabovich, M. P. Pavlou, C. Schiza, E. P. Diamandis, Dynamics of protein expression reveals primary targets and secondary messengers of estrogen receptor alpha signaling in MCF-7 breast cancer cells. Mol Cell Proteomics 15, 2093–2107 (2016).

50. A. Konvalinka, J. Zhou, A. Dimitromanolakis, A. P. Drabovich, F. Fang, S. Gurley, T. Coffman, R. John, S. L. Zhang, E. P. Diamandis, J. W. Scholey, Determination of an angiotensin II-regulated proteome in primary human kidney cells by stable isotope labeling of amino acids in cell culture (SILAC). J Biol Chem 288, 24834–24847 (2013).

51. C. K. Cho, A. P. Drabovich, G. S. Karagiannis, E. Martinez-Morillo, S. Dason, A. Dimitromanolakis, E. P. Diamandis, Quantitative proteomic analysis of amniocytes reveals potentially dysregulated molecular networks in Down syndrome. Clin Proteomics 10, 2 (2013).

52. I. Begcevic, D. Brinc, A. P. Drabovich, I. Batruch, E. P. Diamandis, Identification of brain-enriched proteins in the cerebrospinal fluid proteome by LC-MS/MS profiling and mining of the Human Protein Atlas. Clin Proteomics 13, 11 (2016).

53. I. Begcevic, D. Brinc, L. Dukic, A. M. Simundic, I. Zavoreo, V. Basic Kes, E. Martinez-Morillo, I. Batruch, A. P. Drabovich, E. P. Diamandis, Targeted mass spectrometry-based assays for relative quantification of 30 brain-related proteins and their clinical applications. J Proteome Res 17, 2282–2292 (2018).

54. E. Martinez-Morillo, C. K. Cho, A. P. Drabovich, J. L. Shaw, A. Soosaipillai, E. P. Diamandis, Development of a multiplex selected reaction monitoring assay for quantification of biochemical markers of down syndrome in amniotic fluid samples. J Proteome Res 11, 3880–3887 (2012).

55. C. K. Cho, A. P. Drabovich, I. Batruch, E. P. Diamandis, Verification of a biomarker discovery approach for detection of Down syndrome in amniotic fluid via multiplex selected reaction monitoring (SRM) assay. J Proteomics 74, 2052–2059 (2011).

56. A. Konvalinka, I. Batruch, T. Tokar, A. Dimitromanolakis, S. Reid, X. Song, Y. Pei, A. P. Drabovich, E. P. Diamandis, I. Jurisica, J. W. Scholey, Quantification of angiotensin II-regulated proteins in urine of patients with polycystic and other chronic kidney diseases by selected reaction monitoring. Clin Proteomics 13, 16 (2016).

57. E. Martinez-Morillo, H. M. Nielsen, I. Batruch, A. P. Drabovich, I. Begcevic, M. F. Lopez, L. Minthon, G. Bu, N. Mattsson, E. Portelius, O. Hansson, E. P. Diamandis, Assessment of peptide chemical modifications on the development of an accurate and precise multiplex selected reaction monitoring assay for apolipoprotein e isoforms. J Proteome Res 13, 1077–1087 (2014).

58. A. P. Drabovich, E. P. Diamandis, Combinatorial peptide libraries facilitate development of multiple reaction monitoring assays for low-abundance proteins. J Proteome Res 9, 1236–1245 (2010).

59. Z. Fu, Y. Rais, D. Dara, D. Jackson, A. P. Drabovich, Rational Design and Development of SARS-CoV-2 Serological Diagnostics by Immunoprecipitation-Targeted Proteomics. Anal Chem, (2022).

60. M. Ikawa, N. Inoue, A. M. Benham, M. Okabe, Fertilization: a sperm’s journey to and interaction with the oocyte. J Clin Invest 120, 984–994 (2010).

61. M. S. Chen, K. S. Tung, S. A. Coonrod, Y. Takahashi, D. Bigler, A. Chang, Y. Yamashita, P. W. Kincade, J. C. Herr, J. M. White, Role of the integrin-associated protein CD9 in binding between sperm ADAM 2 and the egg integrin alpha6beta1: implications for murine fertilization. Proc Natl Acad Sci U S A 96, 11830–11835 (1999).

62. A. Jegou, A. Ziyyat, V. Barraud-Lange, E. Perez, J. P. Wolf, F. Pincet, C. Gourier, CD9 tetraspanin generates fusion competent sites on the egg membrane for mammalian fertilization. Proc Natl Acad Sci U S A 108, 10946–10951 (2011).

63. E. Bianchi, B. Doe, D. Goulding, G. J. Wright, Juno is the egg Izumo receptor and is essential for mammalian fertilization. Nature 508, 483–487 (2014).

64. H. Aydin, A. Sultana, S. Li, A. Thavalingam, J. E. Lee, Molecular architecture of the human sperm IZUMO1 and egg JUNO fertilization complex. Nature 534, 562–565 (2016).

65. N. Inoue, Y. Hagihara, D. Wright, T. Suzuki, I. Wada, Oocyte-triggered dimerization of sperm IZUMO1 promotes sperm-egg fusion in mice. Nat Commun 6, 8858 (2015).

66. R. J. Aitken, M. A. Baker, G. F. Doncel, M. M. Matzuk, C. K. Mauck, M. J. Harper, As the world grows: contraception in the 21st century. J Clin Invest 118, 1330–1343 (2008).

67. A. Prakash, T. Rezai, B. Krastins, D. Sarracino, M. Athanas, P. Russo, M. M. Ross, H. Zhang, Y. Tian, V. Kulasingam, A. P. Drabovich, C. Smith, I. Batruch, L. Liotta, E. Petricoin, E. P. Diamandis, D. W. Chan, M. F. Lopez, Platform for establishing interlaboratory reproducibility of selected reaction monitoring-based mass spectrometry peptide assays. J Proteome Res 9, 6678–6688 (2010).

68. A. Prakash, T. Rezai, B. Krastins, D. Sarracino, M. Athanas, P. Russo, H. Zhang, Y. Tian, Y. Li, V. Kulasingam, A. Drabovich, C. R. Smith, I. Batruch, P. E. Oran, C. Fredolini, A. Luchini, L. Liotta, E. Petricoin, E. P. Diamandis, D. W. Chan, R. Nelson, M. F. Lopez, Interlaboratory reproducibility of selective reaction monitoring assays using multiple upfront analyte enrichment strategies. J Proteome Res 11, 3986–3995 (2012).

69. T. D. Karakosta, A. Soosaipillai, E. P. Diamandis, I. Batruch, A. P. Drabovich, Quantification of human kallikrein-related peptidases in biological fluids by multiplatform targeted mass spectrometry assays. Mol Cell Proteomics 15, 2863–2876 (2016).

70. D. Korbakis, D. Brinc, C. Schiza, A. Soosaipillai, K. Jarvi, A. P. Drabovich, E. P. Diamandis, Immunocapture-Selected Reaction Monitoring Screening Facilitates the Development of ELISA for the Measurement of Native TEX101 in Biological Fluids. Mol Cell Proteomics 14, 1517–1526 (2015).

