## Supplementary Figures for "Germ cell-specific proteins ACRV1 and AKAP4 facilitate identification of rare spermatozoa in semen of non-obstructive azoospermia patients"

**List of Supplementary Figures**

Fig. S1. SRM analysis of the matched pre-vasectomy (N=18) and post-vasectomy (N=18) SP to access testis/epididymis specificity of candidate proteins.

Fig. S2. SRM quantification of candidate proteins in SP of pre-vasectomy, post-vasectomy and NOA patients.

Fig. S3. SRM quantification of candidate proteins in SP of NOA patients with the known mTESE outcomes.

Fig. S4. Localization of ADA20_HUMAN and ADA29_HUMAN proteins in motile spermatozoa.

Fig. S5. Imaging flow cytometry identification and visualization of the morphologically normal and intact AKAP4$^+$/ASPX$^+$/Hoechst$^+$ spermatozoa in semen pellet of a normozoospermic patient.

Fig. S6. Imaging flow cytometry identification and visualization of intact AKAP4$^+$/ASPX$^+$/Hoechst$^+$ spermatozoa in semen pellet of a patient diagnosed with oligospermia.

Fig. S7. Visualization of two intact AKAP4$^+$/ASPX$^+$/Hoechst$^+$ spermatozoa in NOA semen pellet.

### Supplementary Figures

#### Figure S1

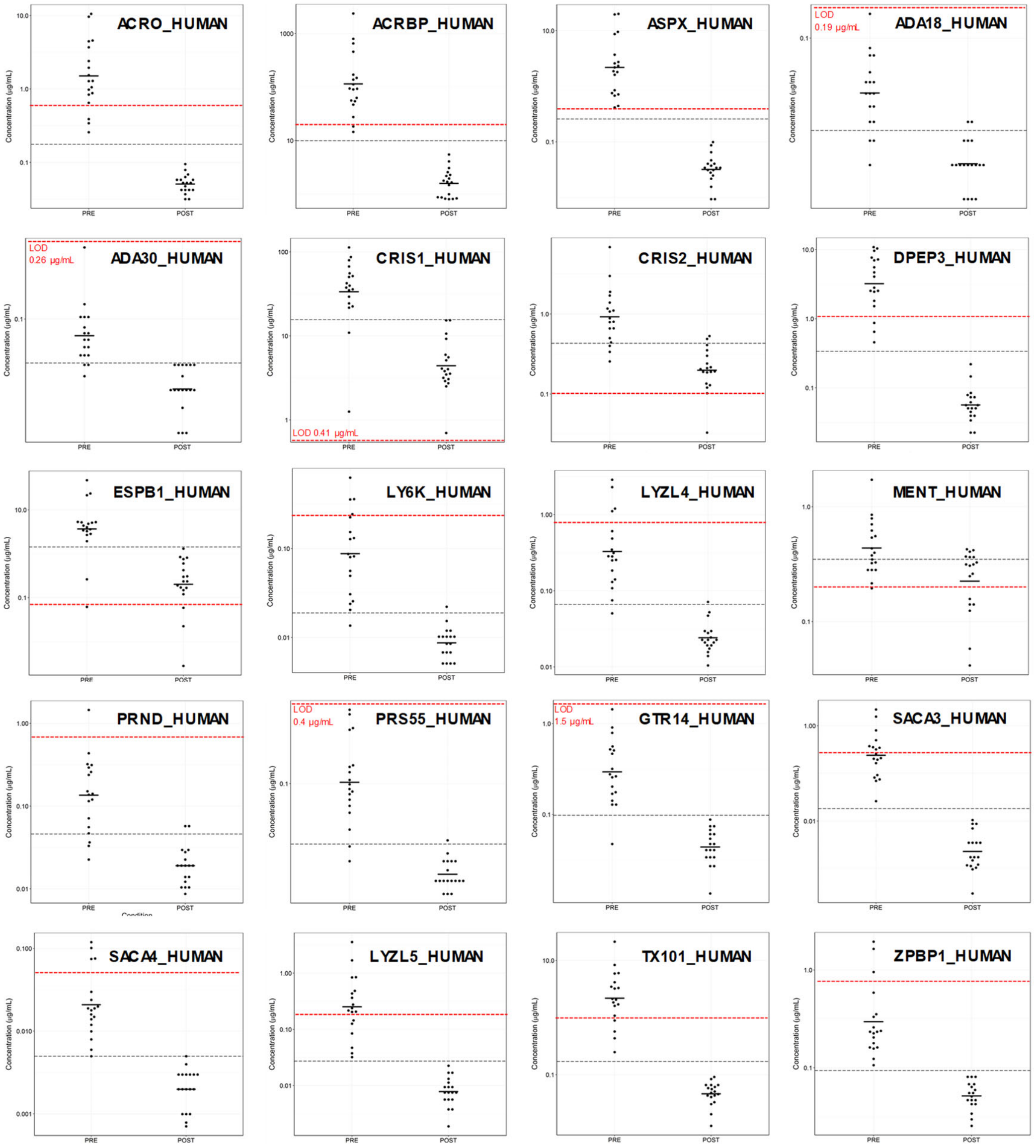


**Fig. S1. SRM analysis of the matched pre-vasectomy (N=18) and post-vasectomy (N=18) SP to access testis/epididymis specificity of candidate proteins.** Grey dashed lines represent apparent cutoffs for the unadjusted raw levels calculated using SRM L/H ratios, while red dashed lines represent SRM assay limits of detections. In post-vasectomy SP, levels of testis- and epididymis-specific proteins decreased below limits of detection. Proteins with the residual expression in prostate or seminal vesicles would still be detectable in some post-vasectomy SP samples (for example, CRIS2_HUMAN and MENT_HUMAN proteins). ASPX_HUMAN at its limit of detection (0.4 ug/mL) revealed ultimate specificity and sensitivity (100%) to detect post-vasectomy.

#### Figure S2

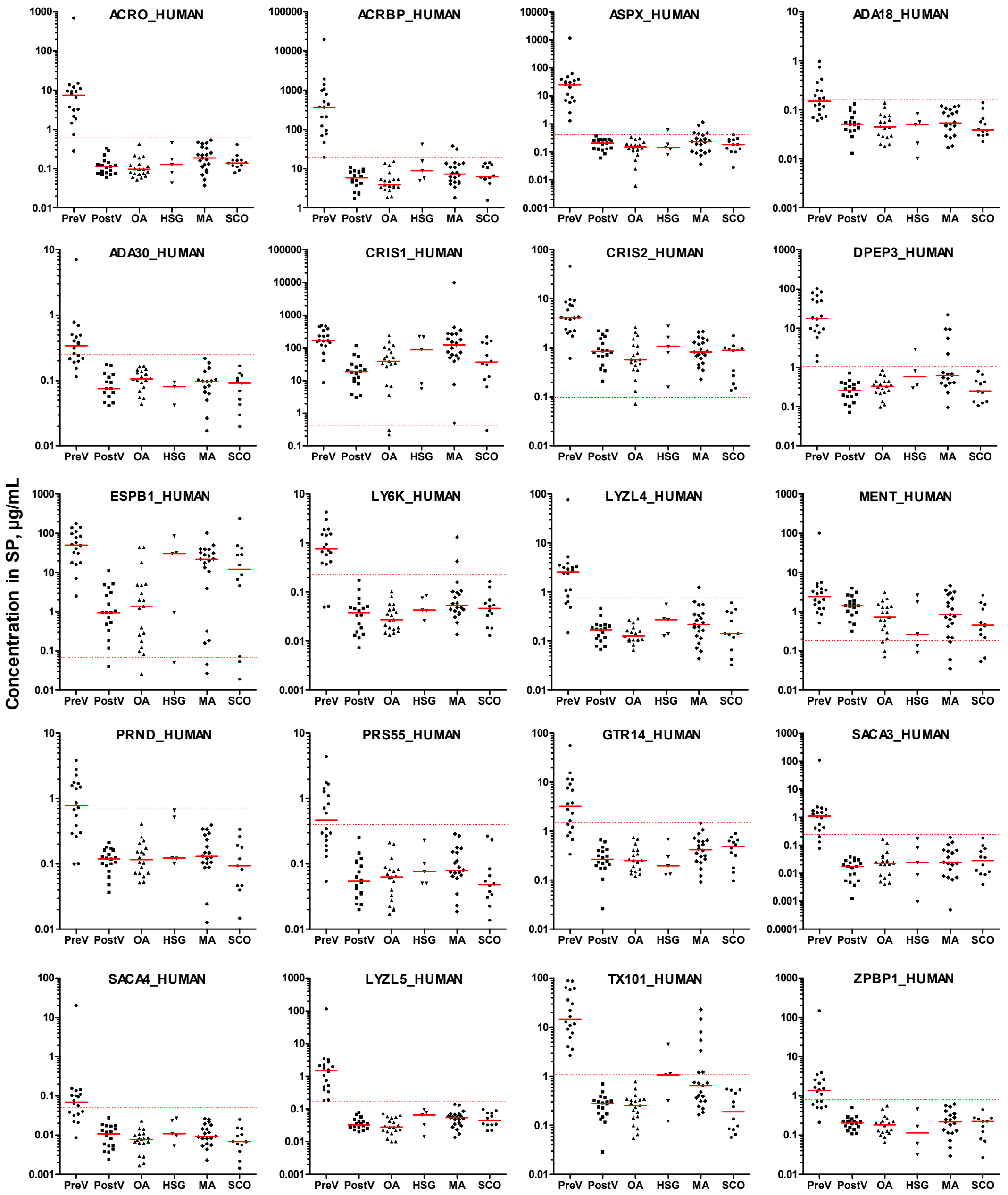


**Fig. S2. SRM quantification of candidate proteins in SP of pre-vasectomy, post-vasectomy and NOA patients.** Testis-specific proteins and two epididymal proteins CRIS1_HUMAN and ESPB1_HUMAN were measured in the independent set of unmatched pre-vasectomy (PreV; N=18) and post-vasectomy (PostV; N=18) SP, as well as SP of OA patients (N=19) and NOA patients with the histological subtypes of hypospermatogenesis (HGS; N=5), maturation arrest (MA; N=21) and Sertoli cell-only (SCO; N=12). Dashed lines represent SRM assay limits of detection.

#### Figure S3

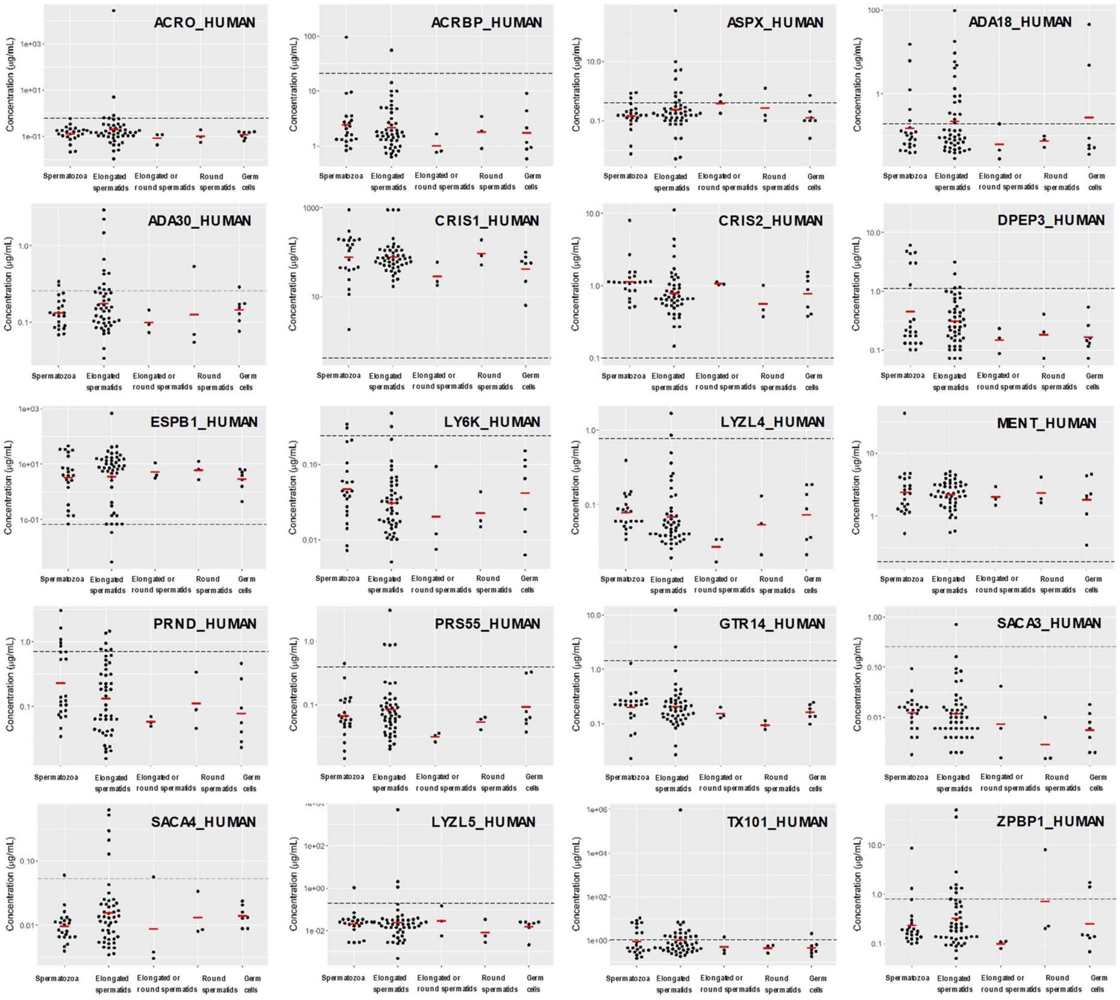


**Fig. S3. SRM quantification of candidate proteins in SP of NOA patients with the known mTESE outcomes.** Testis-specific proteins and two epididymal proteins CRIS1_HUMAN and ESPB1_HUMAN were measured in SP of NOA patients with the known mTESE outcomes and the following cells types extracted with mTESE: spermatozoa (N=22 SP samples), elongated spermatids (N=46), elongated+round spermatids (N=3), round spermatids only (N=3), and immature germ cells (N=7). Dashed lines represent SRM assay limits of detection.

**Figure S4**

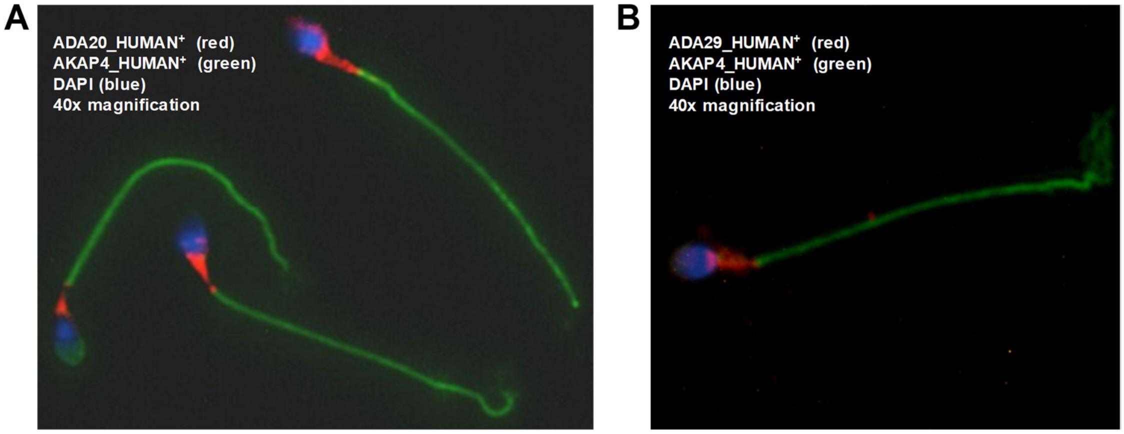


**Fig. S4. Localization of ADA20_HUMAN and ADA29_HUMAN proteins in motile spermatozoa.** Immunofluorescent microscopy analysis of ADA20_HUMAN (ADAM20 gene; red) (**A**) and ADA29_HUMAN (ADAM29 gene; red) (**B**) revealed their expression in the post-acrosomal region of spermatozoa, similar to the testis-specific proteins TX101_HUMAN, DPEP3_HUMAN and LY6K_HUMAN, as we previously reported in Schiza et al. *Mol Cell Proteomics* 2019, 18, 338-351 and *Mol Cell Proteomics* 2018, 17, 2480-2495. Tail and nucleus were stained with AKAP4_HUMAN (green) and DAPI (blue).

**Figure S5**

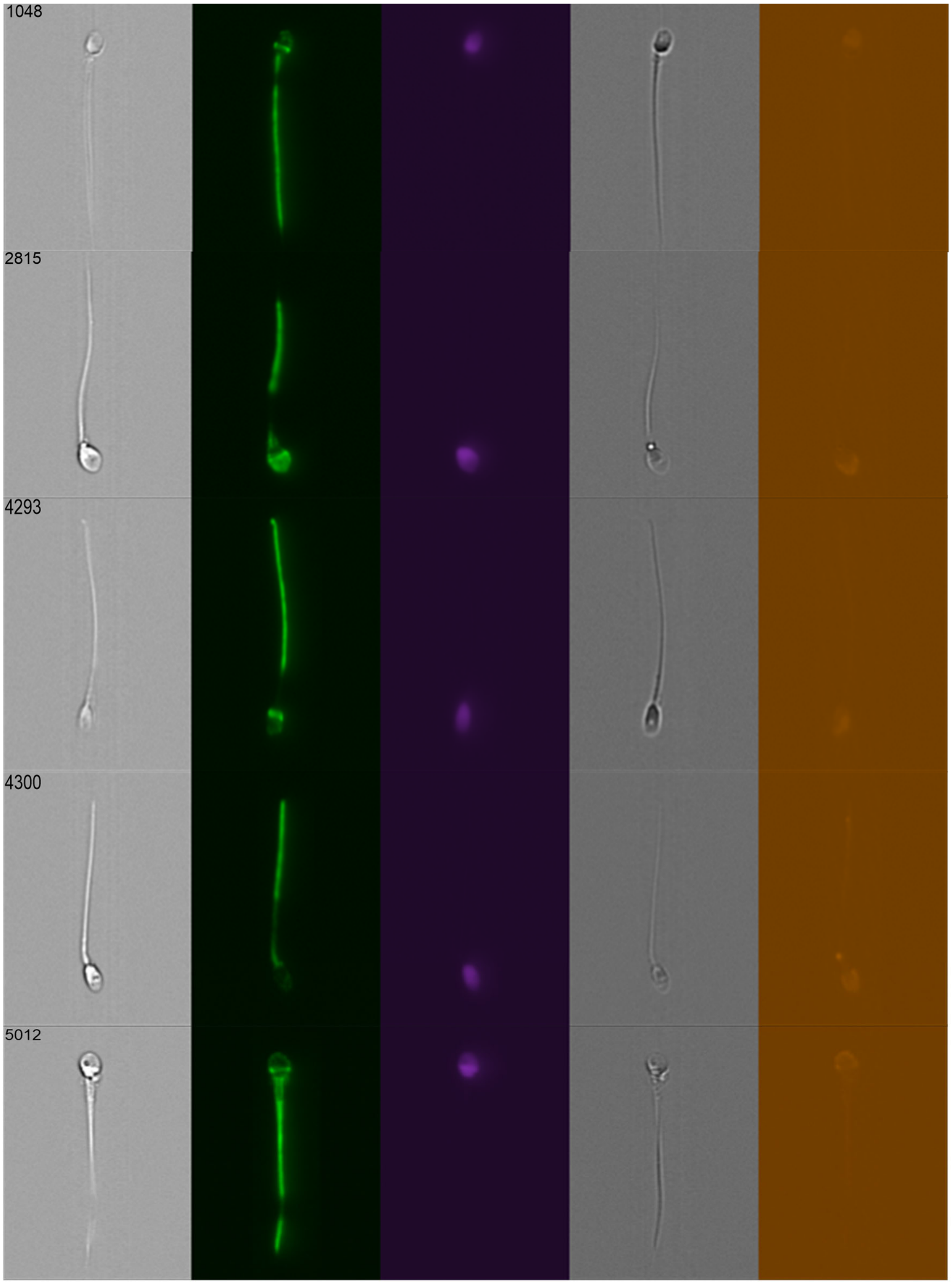


**Fig. S5. Imaging flow cytometry identification and visualization of the morphologically normal and intact AKAP4$^{+}$/ASPX$^{+}$/Hoechst$^{+}$ spermatozoa in semen pellet of a normozoospermic patient.**

**Figure S6**

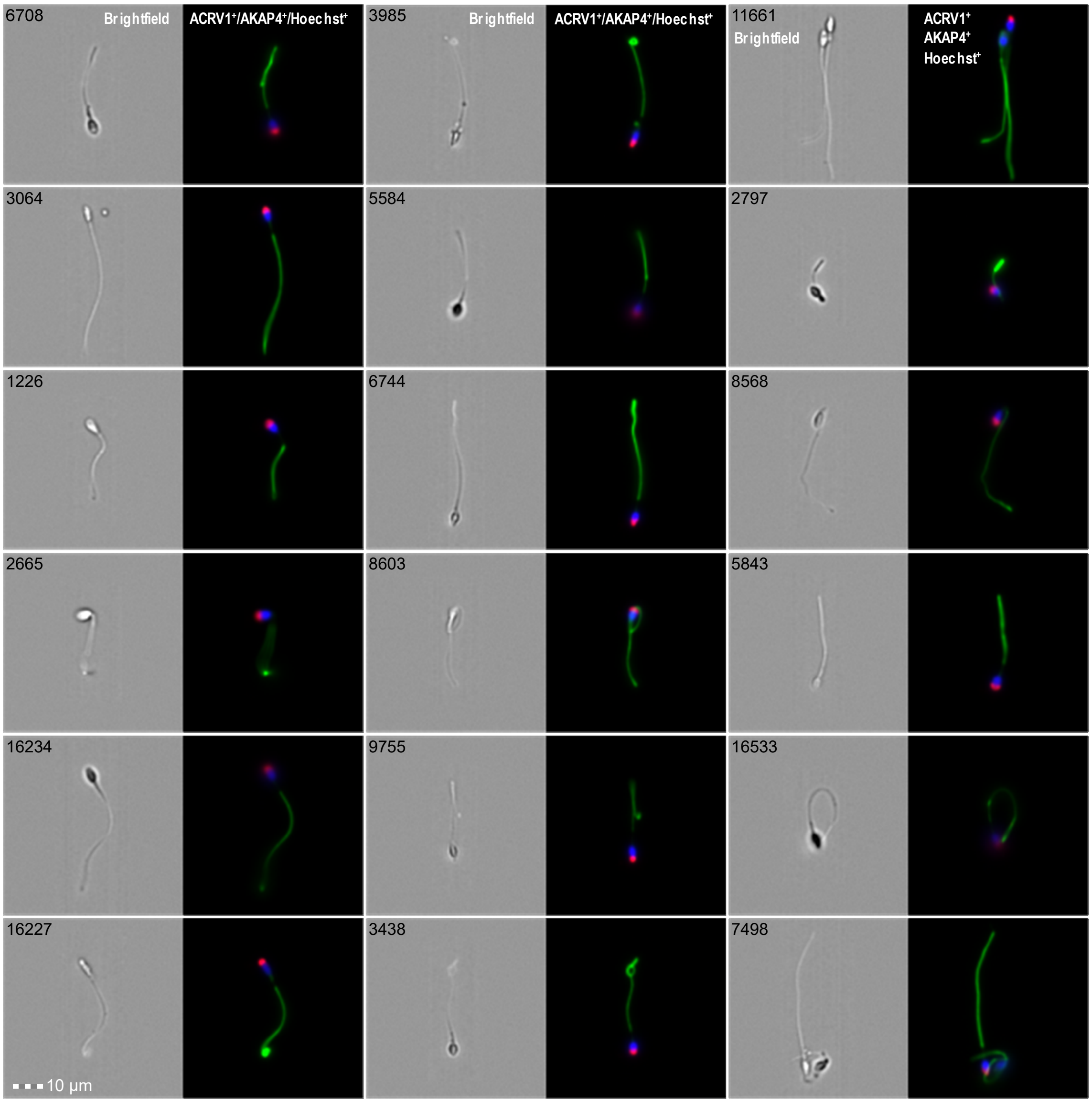


**Fig. S6. Imaging flow cytometry identification and visualization of intact AKAP4$^{+}$/ASPX$^{+}$/Hoechst$^{+}$ spermatozoa in semen pellet of a patient diagnosed with oligospermia.**

#### Figure S7

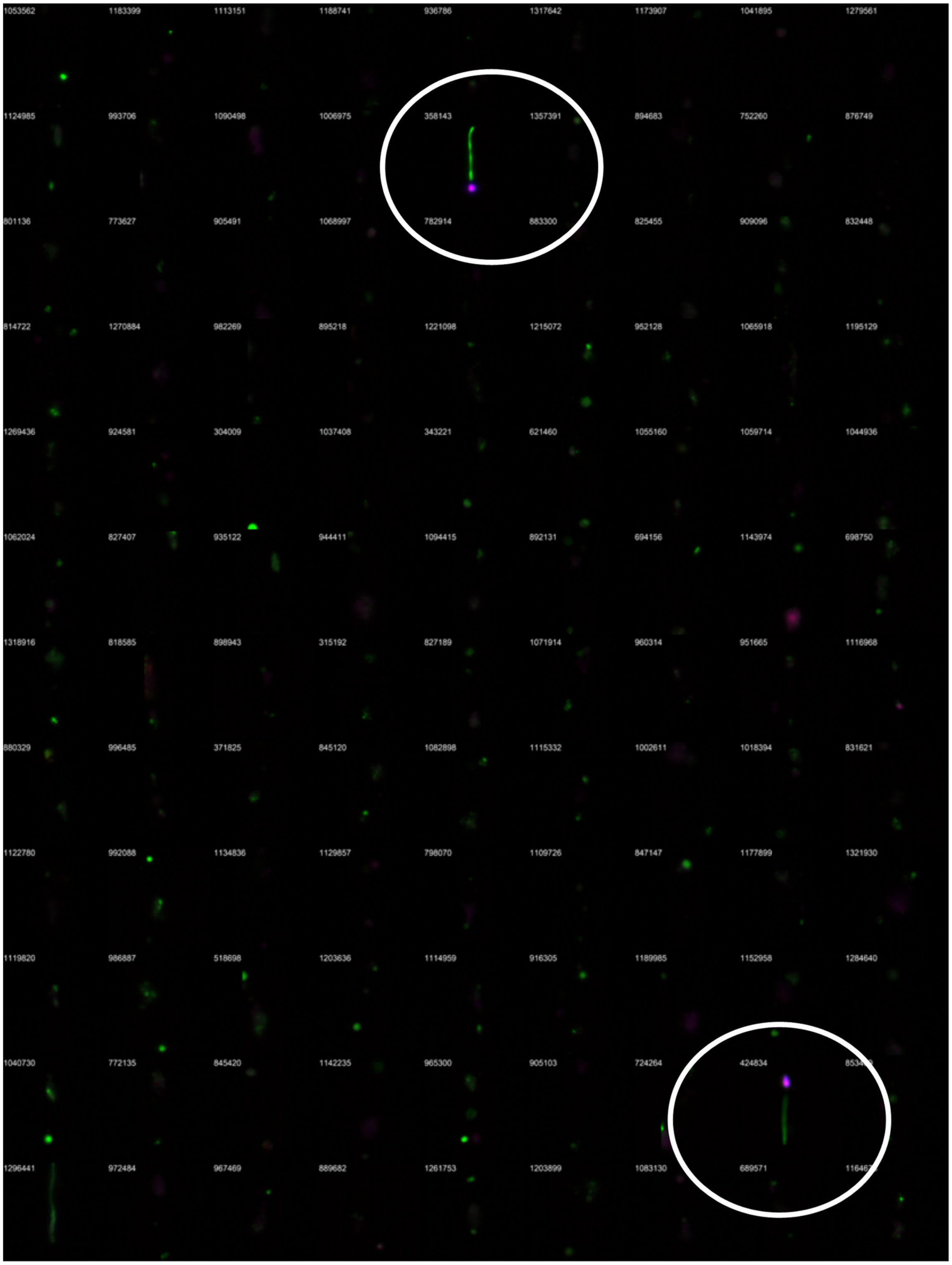


**Fig. S7. Visualization of two intact AKAP4$^+$/ASPX$^+$/Hoechst$^+$ spermatozoa in NOA semen pellet.** The figure includes 108 ImageStream images obtained for the patient #88.
